# Enhanced network synchronization connectivity following transcranial direct current stimulation (tDCS) in bipolar depression: effects on EEG oscillations and deep learning-based predictors of clinical remission

**DOI:** 10.1101/2024.04.19.24306029

**Authors:** Wenyi Xiao, Jijomon C. Moncy, Ali-Reza Ghazi-Noori, Rachel D. Woodham, Hakimeh Rezaei, Elvira Bramon, Philipp Ritter, Michael Bauer, Allan H. Young, Cynthia H.Y. Fu

**Affiliations:** School of Psychology, University of East London, London, UK; Technische Universität Dresden, Dresden, Germany; Department of Psychiatry, University College London, London, UK; Centre for Affective Disorders, Department of Psychological Medicine, Institute of Psychiatry, Psychology and Neuroscience, King’s College London, London, UK; National Institute for Health Research Biomedical Research Centre at South London and Maudsley NHS Foundation Trust, King’s College London, London, UK; South London and Maudsley NHS Foundation Trust, Bethlem Royal Hospital, Beckenham, UK

**Keywords:** transcranial direct current stimulation, brain connectivity, EEG, phase locking value, bipolar disorder, bipolar depression, prediction treatment response

## Abstract

**Aim:** To investigate oscillatory networks in bipolar depression, effects of a home-based tDCS treatment protocol, and potential predictors of clinical response.

**Methods:** 20 participants (14 women) with bipolar disorder, mean age 50.75 ± 10.46 years, in a depressive episode of severe severity (mean Montgomery-Åsberg Rating Scale (MADRS) score 24.60 ± 2.87) received home-based transcranial direct current stimulation (tDCS) treatment for 6 weeks. Clinical remission defined as MADRS score < 10. Resting-state EEG data were acquired at baseline, prior to the start of treatment, and at the end of treatment, using a portable 4-channel EEG device (electrode positions: AF7, AF8, TP9, TP10). EEG band power was extracted for each electrode and phase locking value (PLV) was computed as a functional connectivity measure of phase synchronization. Deep learning was applied to pre-treatment PLV features to examine potential predictors of clinical remission.

**Results:** Following treatment, 11 participants (9 women) attained clinical remission. A significant positive correlation was observed with improvements in depressive symptoms and delta band PLV in frontal and temporoparietal regional channel pairs. An interaction effect in network synchronisation was observed in beta band PLV in temporoparietal regions, in which participants who attained clinical remission showed increased synchronisation following tDCS treatment, which was decreased in participants who did not achieve clinical remission. Main effects of clinical remission status were observed in several PLV bands: clinical remission following tDCS treatment was associated with increased PLV in frontal and temporal regions and in several frequency bands, including delta, theta, alpha and beta, as compared to participants who did not achieve clinical remission. The highest deep learning prediction accuracy 69.45% (sensitivity 71.68%, specificity 66.72%) was obtained from PLV features combined from theta, beta, and gamma bands.

**Conclusions:** tDCS treatment enhances network synchronisation, potentially increasing inhibitory control, which underscores improvement in depressive symptoms. Baseline EEG-based measures might aid predicting clinical response.

## 1. Introduction

Bipolar disorder (BP) is characterized by episodes of elevated mood states as well as depressive and mixed mood states that are associated with changes in sleep and appetite, energy levels, and psychomotor activity (American Psychiatric Association & Association, 2013). Treatment for bipolar depression usually involves medications, such as mood stabilisers and antipsychotic medication, which may be combined with psychotherapy. However, these treatments have limited effectiveness, in part related to individual differences in treatment response and high rates of discontinuation due to intolerability and adverse effects (Chakrabarti, 2014; McIntyre et al., 2022).

The non-invasive brain stimulation, transcranial direct current stimulation (tDCS), is a potential treatment option for bipolar depression (Mutz et al., 2018, 2019, Woodham et al., 2021). tDCS generates a weak direct current (0.5 - 2.0 mA) which is applied to the scalp through electrodes. In bipolar depression, the anode electrode is usually placed over the left dorsolateral prefrontal cortex (DLFPC) and cathode electrode over the right DLPFC, frontotemporal or suborbital region (Tortella et al., 2015; Mutz et al., 2018, 2019). The current modulates resting membrane potential, in which anode stimulation increases neuronal excitability with a reduction in GABAergic activity while cathode stimulation decreases neuronal excitability by reducing glutamate levels (Stagg et al., 2018). tDCS does not lead to neuronal depolarisation, in contrast to repetitive transcranial magnetic stimulation (rTMS), and it does not lead to a generalised seizure, in contrast to electroconvulsive therapy (ECT). tDCS stimulation has been found to modulate functional interactions in brain networks extending beyond the regions directly targeted by stimulation, impacting on the wider neural network underlying mood regulation (Polania et al., 2011; Kunze et al., 2016; Woodham et al., 2021).

The DLPFC is a key region in emotion processing and executive functioning, which are impaired in bipolar depression (Hassel et al., 2008; Townsend et al., 2010). Imbalances in activity in the left and right DLPFC, namely hypoactivity in the left DLPFC and hyperactivity in the right DLPFC, is found in depression (Grimm et al., 2008; Maeda et al., 2000). The prefrontal cortex has a crucial role in regulating responses to threat by directly inhibiting the amygdala complex, in which electrical stimulation over the prefrontal cortex inhibits amygdala response (Quirk et al., 2003).

Oscillations in electrical activity between brain regions range from slow to fast frequencies, which reflect coupling between regions during different states and in mental health disorders. Synchronization in oscillatory networks underlies the variable affective states in bipolar disorder (Chen et al., 2008, Kam et al., 2013). Electroencephalography (EEG) provides a measure of electrical activity in brain regions, generating assessments of the strength and connections between regions.

In bipolar disorder, increased power in high frequency bands, namely beta and gamma, has been observed as compared to healthy participants (Kam et al., 2013). In bipolar depression though, reductions in the high frequency beta/gamma bands have been reported (Canali et al., 2015), and increasing depressive severity was associated with decreased gamma band synchronization (Kim et al., 2013). Following rTMS treatment, clinical response was associated with increased strength in EEG functional connectivity, as measured by beta and gamma phase locking value (PLV) in frontal region and temporal-parietal regions in bipolar depression (Zuchowicz et al., 2019), and increased power in low frequency delta and theta bands at baseline was associated with a subsequent clinical response to rTMS in unipolar and bipolar depression (Woźniak-Kwaśniewska et al., 2015). However, oscillatory network synchronization following tDCS treatment has not been examined in bipolar depression. Furthermore, by applying data-driven artificial intelligence algorithms, we may be able to develop predictors of clinical response (Fu and Costafreda, 2013).

In the present study, we sought to investigate the oscillatory networks in bipolar depression, effects of a home-based tDCS treatment protocol, and potential predictors of clinical response. We utilized a portable wireless EEG device with 4-dry electrodes which has demonstrated robust signal properties (Cannard et al., 2021; Krigolson et al., 2021), participants underwent EEG acquisition in their own homes with real-time supervision by video conference. We investigated EEG metrics, namely power and PLV, associated with changes in depressive severity as well as the categorial effects of clinical remission. To explore preliminary EEG-based predictors, we applied deep learning to examine whether baseline EEG measures could predict clinical remission. Based on previous findings in rTMS treatment (Woźniak-Kwaśniewska et al., 2015), we expected that oscillations in low frequency bands might be predictive of treatment response.

## 2. Methods

### 2.1. Participants

Ethical approval was provided by London Fulham Research Ethics Committee. All participants provided written informed consent electronically. The study was an open-label, single arm acceptability and feasibility trial of home-based tDCS treatment for bipolar depression (Ghazi-Noori et al., 2024). Participants were aged 18 years or above, with a diagnosis of bipolar disorder and in a current major depressive episode without psychotic features, based on Diagnostic and Statistical Manual of Mental Disorders, Fifth Edition (DSM-5) (American Psychiatric Association & Association, 2013), determined by a structured assessment using the Mini-International Neuropsychiatric Interview (MINI; Version 7.2) (Sheehan et al., 1998). All participants had at least a moderate severity of depressive symptoms, as measured by a minimum score of 18 on the Montgomery-Åsberg Depression Rating Scale (MADRS) (Montgomery and Åsberg 1979). Participants were either medication-free or taking a stable regime of mood-stabilizing medication or in psychotherapy for a minimum of two weeks. Exclusion criteria included significant suicide risk, comorbid psychiatric disorder, and contraindications to tDCS. Ghazi-Noori et al. (2024) provides a full description of the study.

### 2.2. tDCS treatment protocol

The protocol consisted of a 6-week course of active tDCS, which was self-administered by participants in their homes 5 times a week for 3 weeks and then twice a week for 3 weeks, for a total of 21 sessions. A member of the research team was present at each session by Microsoft Teams video call. Clinical remission was defined as the MADRS score of less than 10 at the end of the treatment.

A bifrontal montage was applied with the anode positioned over left DLPFC (F3 position according to international 10/20 EEG system) and cathode over right DLPFC (F4 position). Each electrode was a 23cm^2^ conductive rubber electrode covered by saline soaked sponges. Simulation was 2 mA for a duration of 30 minutes with a gradual ramp up over 120 seconds at the start and ramp down over 15 seconds at the end of each session. The Flow Neuroscience tDCS device was used for all participants.

### 2.3. Remote EEG acquisition and preprocessing

EEG data were acquired at two time points: at baseline, prior to the start of treatment (pre-treatment), and at the end of treatment (post-treatment). EEG data had been acquired in a sub-sample of 22 participants (15 women; mean age 51.59 years) at baseline, however, data were not available for one participant post-treatment and data from another participant was of poor quality and was not included. Thus, data were available in 20 participants (14 women; mean age 50.75 years, SD10.46 years), at pre-treatment and post-treatment (Table 1).

**Table 1.** Demographic characteristics of participants. Categorical variables are presented as number of participants with percentage in parentheses for treatments during trial. Duration current depressive episode is presented as mean with range in parentheses. Mean values are presented with ‘±’ standard deviation values. MADRS, Montgomery-Åsberg Depression Rating Scale. There is no significant difference between remission and non-remission group at age (*t* = -1.896, *p* = 0.077) or gender (*χ*^2^ = 0.616, *p* = 0.433). However, there is a significant difference at the baseline MADRS score (*t* = -2.365, *p* = 0.030), and at week 6 post-treatment (*t* = -5.429, *p* < 0.001).

| Characteristic | Remission | Non-Remission |
| --- | --- | --- |
| Total number (women) | 11 (9) | 9 (5) |
| Age | 47.19 ± 12.15 | 55.11 ± 6.05 |
| Years of education | 17.82 ± 1.94 | 15.67 ± 2.18 |
| IQ | 102.55 ± 9.17 | 102.89 ± 7.62 |
| Duration current depressive episode (weeks) | 54.45 (3-260) | 92.56 (8-64) |
| MADRS score at baseline | 24.00 ± 3.13 | 26.89 ± 2.15 |
| MADRS score at end of treatment | 5.18 ± 1.72 | 14.40 ± 4.88 |
| Treatments during trial |  |  |
| Taking mood stabilizer and other medications | 10 | 6 |
| Taking antidepressant medication only | 0 | 1 |
| Taking no medication | 1 | 2 |
| Engaged in psychotherapy | 4 | 1 |

At each EEG acquisition session, a trained research team member provided real-time guidance via videoconference. At each EEG session, four 5-minute EEG recordings were acquired. Each recording measured at resting state in which participants were asked to maintain a relaxed posture without any body movements. The resting state recordings were conducted in the following order: eyes closed, eyes open, eyes closed, and eyes open. The two five-minutes resting state eyes-closed recording at pre- and post-treatment were used the present analysis.

EEG recordings were acquired using a Muse device, a wireless EEG device equipped with 4 dry electrodes (Figure 1). Sampling frequency was 256 Hz. Frontal electrode positions were AF7 and AF8, and the temporoparietal positions were TP9 and TP10. EEG signals were referenced to the FPz electrode.

**Figure 1.**
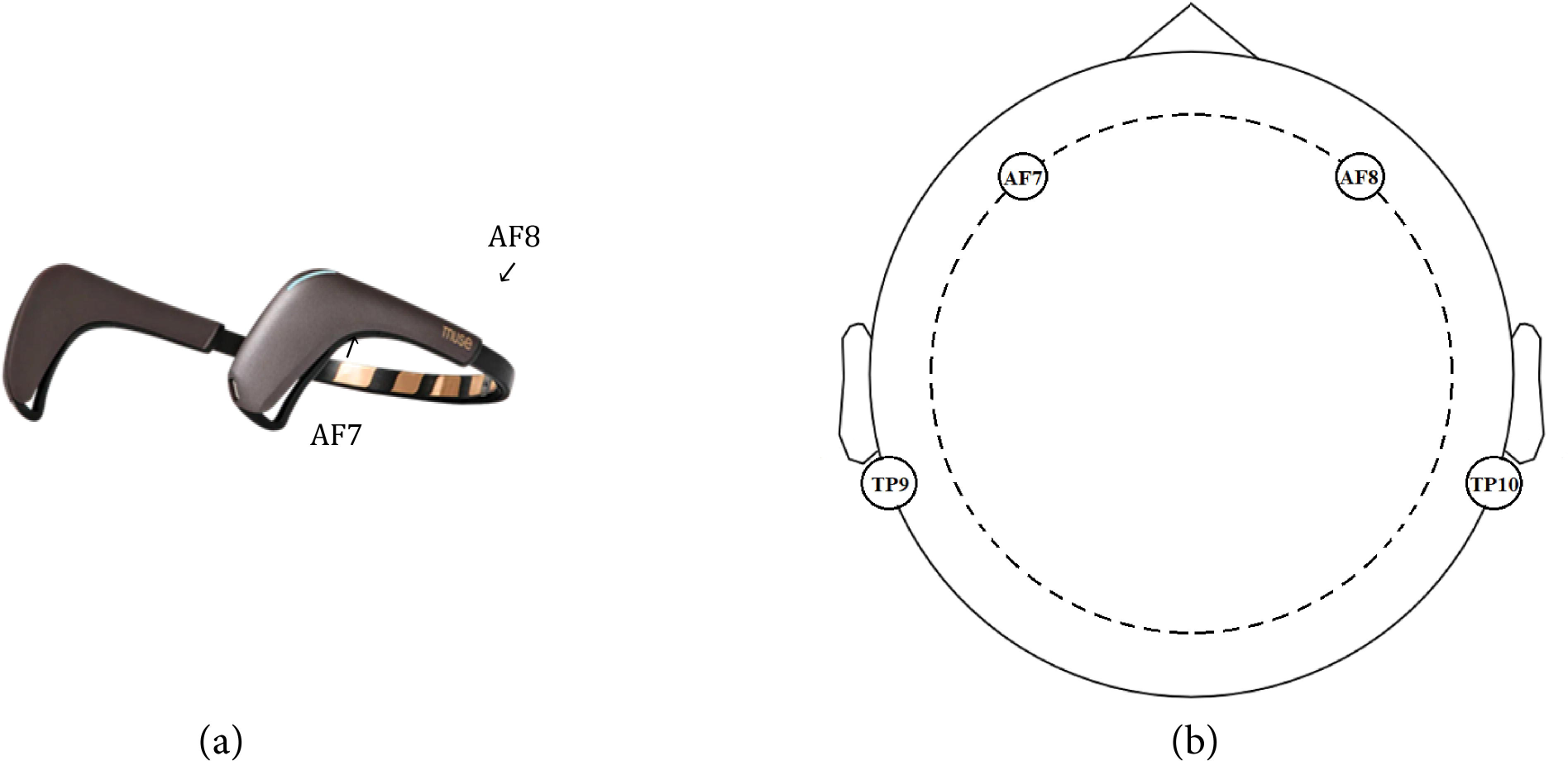
Image of (a) EEG device (Muse), which consisted of four electrode (AF7, AF8, TP9 and TP10), and (b) EEG distribution of electrodes based on international 10/20 positioning.

Each recording was segmented into 60 separate EEG windows, each lasting 10 seconds without overlap. The recorded EEG signals, saved in CSV format, included timestamps for each EEG sample, raw EEG signals from each electrode, Horse Shoe Indicator (HSI) values for each electrode. HSI values served as indicators of electrode connectivity quality: HSI value 1 indicates excellent connectivity between electrode and the participant’s scalp, 2 indicates average connectivity, and 4 poor connectivity. Values were averaged across the samples in each window, and all windows with an average HSI of 2 or less were selected.

Each EEG signal was segmented into 10-second windows. EEG signals from each electrode undergo filtering across six distinct frequency bands: the full band (1-60 Hz), delta δ (1-4 Hz), theta θ (4-8 Hz), alpha α (8-12 Hz), beta *β* (12-30 Hz), and gamma *γ* (30-60 Hz). Employing Butterworth Infinite Impulse Response (IIR) filters of 5th order, the signals are effectively filtered.

### 2.4. EEG analysis

EEG band power and phase locking value (PLV) were extracted as resting state EEG metrics. EEG band power was calculated for all four electrodes (AF7, AF8, TP9, TP10). PLV was computed for all possible electrode pairs (AF7-AF8, AF7-TP9, AF7-TP10, AF8-TP9, AF8-TP10, TP9-TP10). PLV assesses the phase synchrony between two time series signals (Hoke et al., 1989; Lachaux et al., 1999). It is a common measure employed to determine functional connectivity between EEG signals recorded from two electrodes, offering insight into the temporal relationships of neural signals independent of their amplitude. PLV is a statistical metric constrained within the range of 0 to 1. PLV value approaching 1 indicates high phase synchronization with minimum variation in phase difference across the EEG signals, while a value close to 0 suggests no phase synchronization. A total of 24 band power values and 36 PLVs were computed with two sets of 60 EEG measurements at pre- and post-treatment, were available for statistical analysis. Full description is presented in Supplementary Materials.

### 2.5. Statistical analyses

Statistical analyses of band power and PLV were performed to investigate potential associations between changes in EEG measures and depression severity and to assess effects of remission and non-remission status following tDCS treatment.

Pearson’s correlation analysis was conducted to examine the relationship between changes in EEG measures and the proportional change in MADRS scores from baseline to the end of the 6-week treatment period, across all participants. The proportional change in MADRS is calculated by subtracting the baseline MADRS score from the MADRS score after 6 weeks of treatment, then dividing this absolute difference by the baseline MADRS score.

Factorial analyses with and without proportional change in MADRS as covariate were used for between-group comparisons. To test whether EEG measures change in response to treatment, a two-way ANOVA was performed for each EEG variable. The factors were: remission group (remission, non-remission) and Time (baseline pre-treatment, post-treatment). A total of 60 statistical tests were performed.

The coefficients were estimated in the R statistical environment (R core team, 2021) using linear regression (*lm* built-in function). Post hoc tests (Tukey honestly significant difference (HSD)) were performed to assess significant effects. The statistical threshold was set at *p*<0.05, with correction for multiple comparisons by controlling False Discovery Rate (FDR). A full description is in the supplementary materials.

### 2.6. Deep learning analysis

Participants were categorized into two groups based on their remission status following tDCS treatment. In the classification analysis, remission was defined as the positive class and non-remission as the negative class, with sensitivity representing remission and specificity representing non-remission. Values were extracted from each EEG frequency band at pre-treatment.

From each EEG band, PLV feature vectors with a dimensionality of 6 are generated, representing each of the six electrode pairs. These six-dimensional feature vectors, both individually and through the concatenation of PLV features from multiple EEG bands, were employed as inputs for deep learning models with varying parameters. This concatenation process at the feature level led to a linear increase in the feature dimension. To assess the effectiveness of different combinations of PLV features from individual bands and combinations of multiple bands, two distinct deep learning architectures were investigated. The first architecture utilized a fully connected perceptron deep learning structure, and the second employed a one-dimensional convolutional neural network (1DCNN) architecture. Considering combination of features from multiple EEG bands, the dimensionality varied as follows: 6 (single band), 12 (two-band combination), 18 (three-band combination), 24 (four-band combination), and 30 (combination of all bands).

For the fully connected perceptron deep learning network, a four-layer architecture was implemented, comprising layers with 32, 32, 16, and 1 perceptrons (output layer) for all input combinations, except in the case of the all-band combination where the feature size was 3. In this scenario, 64 perceptrons were employed in the first layer. The all-band combination encompassed all PLV features extracted from the delta to gamma bands. In the 1DCNN-based architecture, the initial fully connected layer of the perceptron network was replaced with a convolutional layer. This convolutional layer employed a kernel size of 3. To ensure kernel overlap, the number of filters used was determined by multiplying the input dimension by a multiplication factor 2/3. Following the convolutional layer, a MaxPooling1D layer with a pool size of 2 was integrated to downsample the feature maps, aiming to extract the most relevant features while reducing computational complexity. For single-band PLV features with an input dimension of 6, 4 filters with a kernel size of 3 were used. Similarly, for dimensions 12, 18, 24, and 30, 8, 12, 16, and 20 filters were employed respectively. The activation function ‘relu’ was applied to all layers except the output layer, where ‘sigmoid’ was utilized. To minimize the ‘binary cross-entropy’ loss function, the ‘adam’ optimizer was employed.

Due to the constrained size of the dataset and our emphasis on accurately evaluating model performance over computational efficiency, we opted for the Leave-One-Subject-Out (LOSO) methodology to assess the deep learning model’s accuracy. Employing this approach, we conducted 20 iterations of training and testing for each input combination. During each iteration, one participant out of the 20 was reserved for testing, while the PLV features from the remaining 19 participants were utilized for model training. Following 50 epochs of model training, we identified the most effective model based on its classification accuracy on a validation set. This validation set, comprising 240 randomly selected vectors, equally distributed between remission and non-remission groups, was drawn from the training data. The model exhibiting the highest classification accuracy on the validation set underwent further testing.

## 3. Results

### 3.1. Clinical outcome

Following tDCS treatment, 11 participants attained clinical remission (mean MADRS score post-treatment 5.18, SD 1.72) and 9 participants were in non-remission (mean MADRS score post-treatment 14.40, SD 4.88), which was a significant difference (*t* = -5.429, *p* < 0.001).

### 3.2. Relationship between changes in depression severity and EEG power

No significant correlation was found between change in EEG power and proportional change in MADRS scores from pre-to post-treatment.

### 3.3. Relationship between changes in depression severity and EEG PLV connectivity

Positive correlations with an improvement in depressive symptoms following tDCS treatment were found in delta band PLV in several electrode pairs: AF7-AF8 (*R^2^ = 0.48,* FDR-adjusted *p* = 0.032), AF7-TP9 (*R^2^ = 0.55,* FDR-adjusted *p* = 0.013), AF8-TP9 (*R^2^ = 0.56*, FDR-adjusted *p* = 0.001), and TP9-TP10 (*R^2^ = 0.49,* FDR-adjusted *p* = 0.027), as well as in beta PLV in pair: AF8-TP10 (*R^2^ = 0.45,* FDR-adjusted *p* = 0.044) (Figures 3, Supplementary Table 5). No regions showed a negative correlation with an improvement in depressive symptoms.

**Figure 2.**
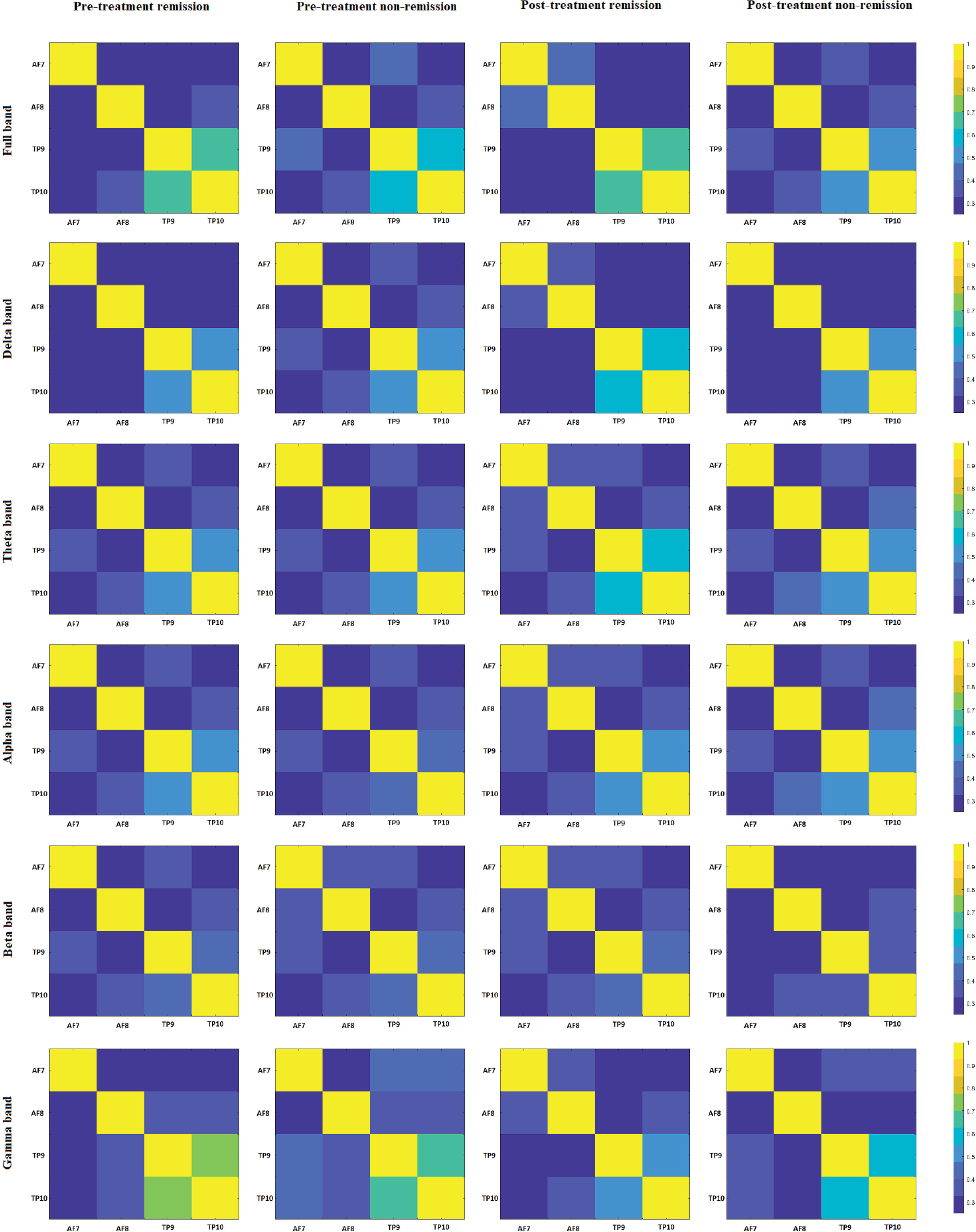
Image representing averaged PLV values across different group (remission and non-remission group) and time (pre- and post-treatment). Rows depict EEG bands: full band, delta, theta, alpha, beta and gamma, and columns represent participant categories: pre- treatment remission, pre-treatment non-remission, post-treatment remission, and post- treatment non-remission groups.

**Figure 3.**
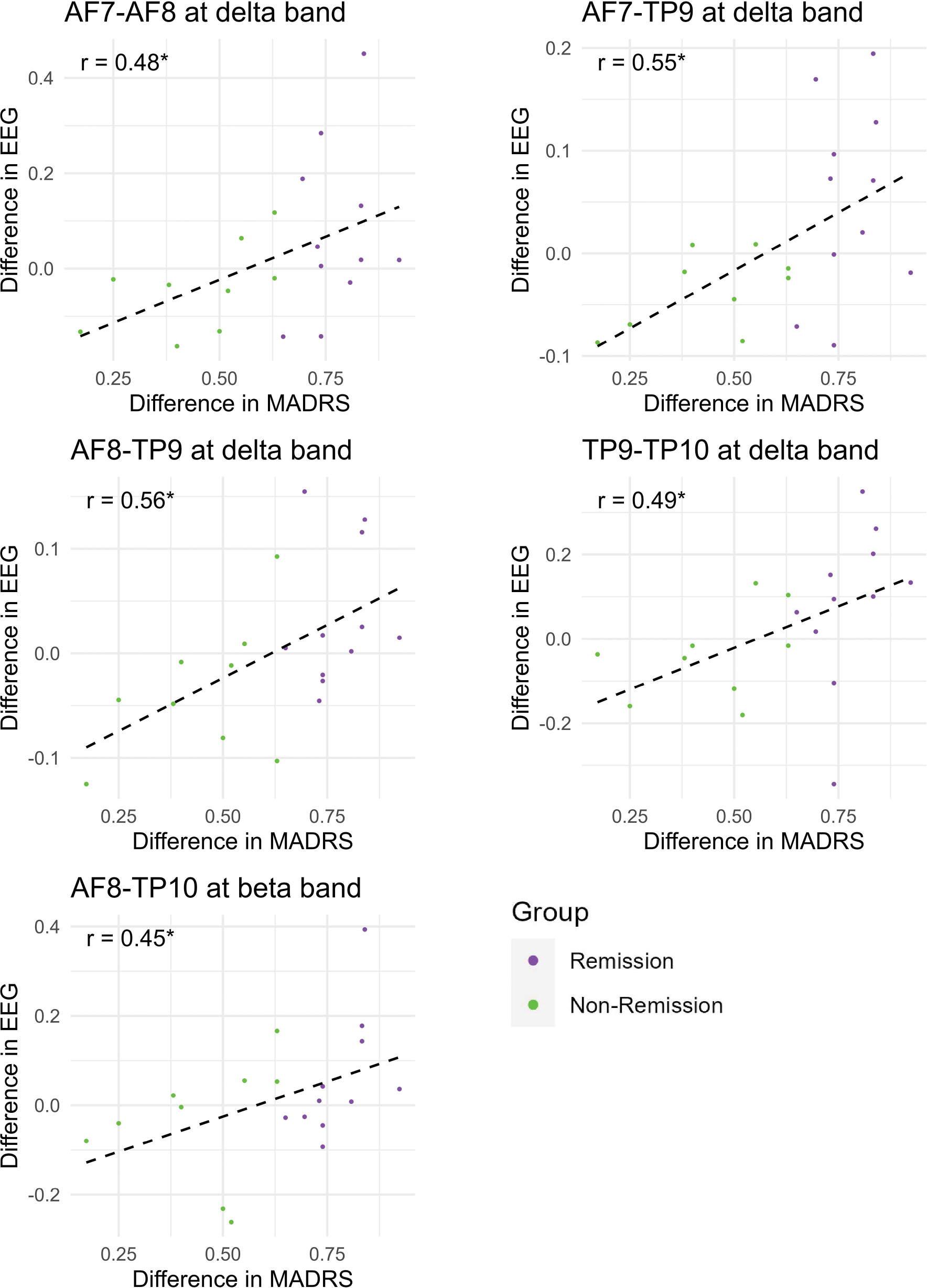
Scatter plots of phase locking value (PLV) (y-axis: Difference in EEG) of variables showing significant positive correlation with proportional change in MADRS (x-axis: Difference in MADRS) across participants. Positive correlations were observed for each PLV synchronization EEG channel pair.

### 3.4. Effects of remission status in EEG power

No significant differences in power for each frequency band before and after stimulation were found for any of groups.

### 3.5. Effects of remission status in EEG PLV connectivity

A significant main effect of group was observed in whole band PLV in AF7-TP9 (*F* = 5.762, FDR-adjusted *p* = 0.022). When examining the indices for the five frequency bands separately, significant main effects of group were observed in PLV in the following channel pairs: in delta band: AF7-AF8 (*F* = 8.874, FDR-adjusted *p* = 0.005), AF7-TP10 (*F* = 6.939, FDR-adjusted *p* = 0.012), AF8-TP9 (*F* = 4.346, FDR-adjusted *p* = 0.044) and TP9-TP10 (*F* = 5.62, FDR-adjusted *p* = 0.023); in theta band: AF7-AF8 (*F* = 11.917, FDR-adjusted *p* = 0.001), TP9-TP10 (*F* = 8.12, FDR-adjusted *p* = 0.007); in alpha band: AF7-AF8 (*F* = 9.777, FDR-adjusted *p* = 0.003), AF7-TP9 (*F* = 5.393, FDR-adjusted *p* = 0.026), AF7-TP10 (*F* = 8.515, FDR-adjusted *p* = 0.006) and TP9-TP10 (*F* = 8.032, FDR-adjusted *p* = 0.007); in beta band: AF7-TP9 (*F* = 8.322, FDR-adjusted *p* = 0.007), AF7-TP10 (*F* = 7.235, FDR-adjusted *p* = 0.011) (Supplementary Table 1). Post-hoc tests demonstrated that for each electrode pair, remission group showed a significantly higher PLV as compared to non-remission group (Figure 4, Supplementary Table 2).

**Figure 4.**
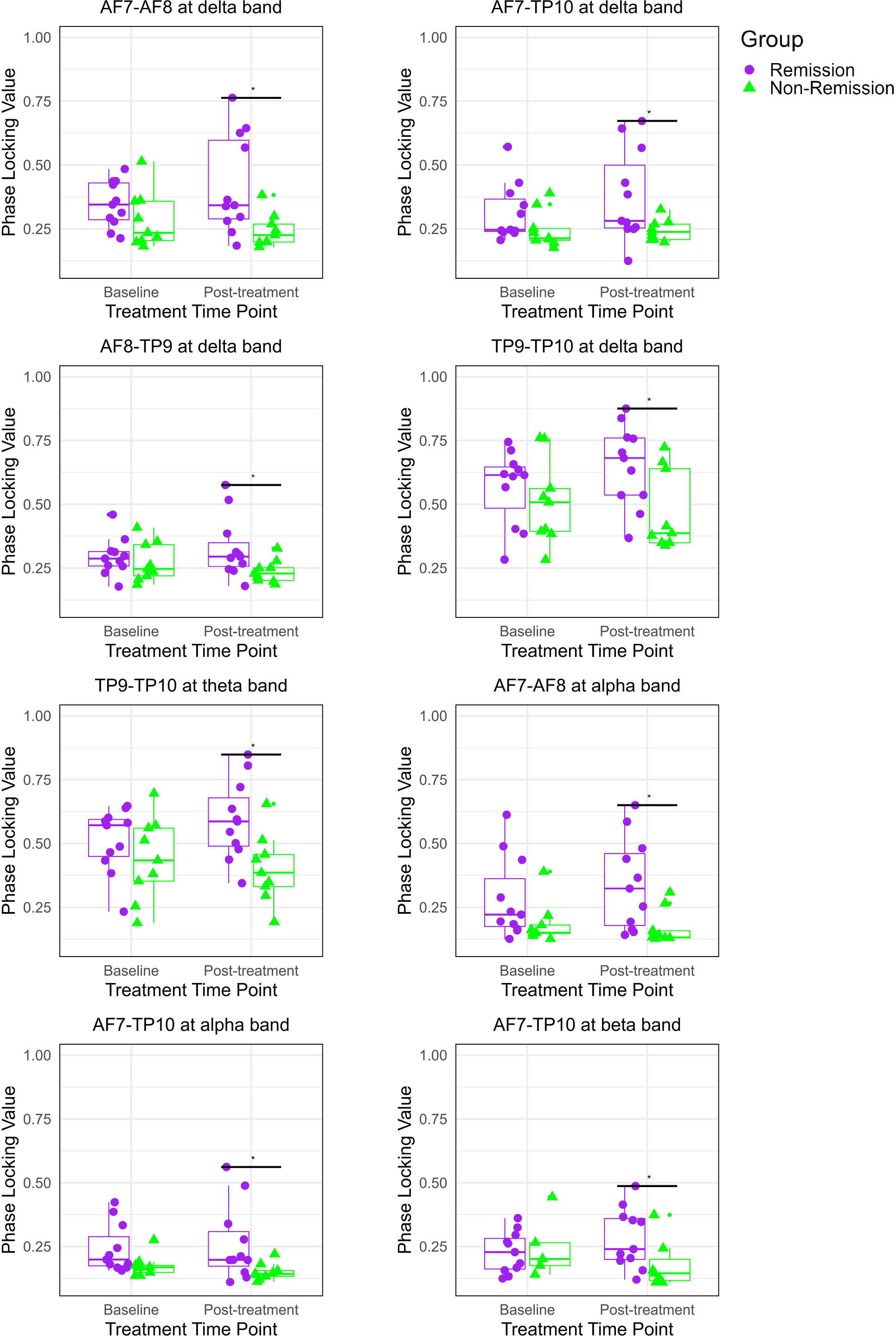
Boxplots comparing phase locking values (PLV) between remission and non-remission groups within specific frequency bands. The remission group exhibited increased PLV as compared to the non-remission group post-treatment across electrode pairs within delta, theta, alpha and gamma frequency ranges. Significant differences are denoted by *, indicating adjusted *p*-values corrected for false discovery rate (FDR) below 0.05. Purple colour signifies the remission group, and green indicates the non-remission group. Significant main effects for groups in the PLV of AF7-TP9 were found across the full, alpha, and beta bands, in which post-hoc tests showed significant differences between groups at post-treatment. Remission group showed increased PLV value as compared to the non-remission group at post-treatment, including PLV of AF7 and AF8 in both delta and alpha bands (*t_delta_* = 2.88, FDR-adjusted *p* = 0.01; *t_alpha_* = 2.93, FDR-adjusted *p* = .001); PLV of AF7 and TP10 in delta and beta bands (*t_delta_* = 2.34, FDR-adjusted *p* = 0.04; *t_beta_* = 2.60, FDR-adjusted *p* = 0.02); PLV of TP9 and TP10 in delta and theta bands (*t_delta_*= 2.51, FDR-adjusted *p* = 0.02; *t_theta_* = 2.93, FDR-adjusted *p* = 0.01) and PLV of AF8 and TP9 in the delta band (*t* = 2.32, FDR-adjusted *p* = .04).

A significant main effect of time was found in the gamma band: PLV AF8-TP9 (*F* = 6.138, FDR-adjusted *p* = 0.018), and TP9-TP10 (*F* = 4.766, FDR-adjusted *p* = 0.036), in which there was increased PLV at baseline pre-treatment as compared to post-treatment (Supplementary Table 1)

A significant interaction effect of remission group by time was found in the beta band PLV TP9-TP10 (*F* = 4.48, FDR-adjusted *p* = 0.041), in which there were no differences between the remission and non-remission groups at baseline, however remission group showed an increase in PLV from baseline to post-treatment while the non-remission group showed a significant decrease from baseline to post-treatment (Figure 5, Supplementary Table 1).

**Figure 5.**
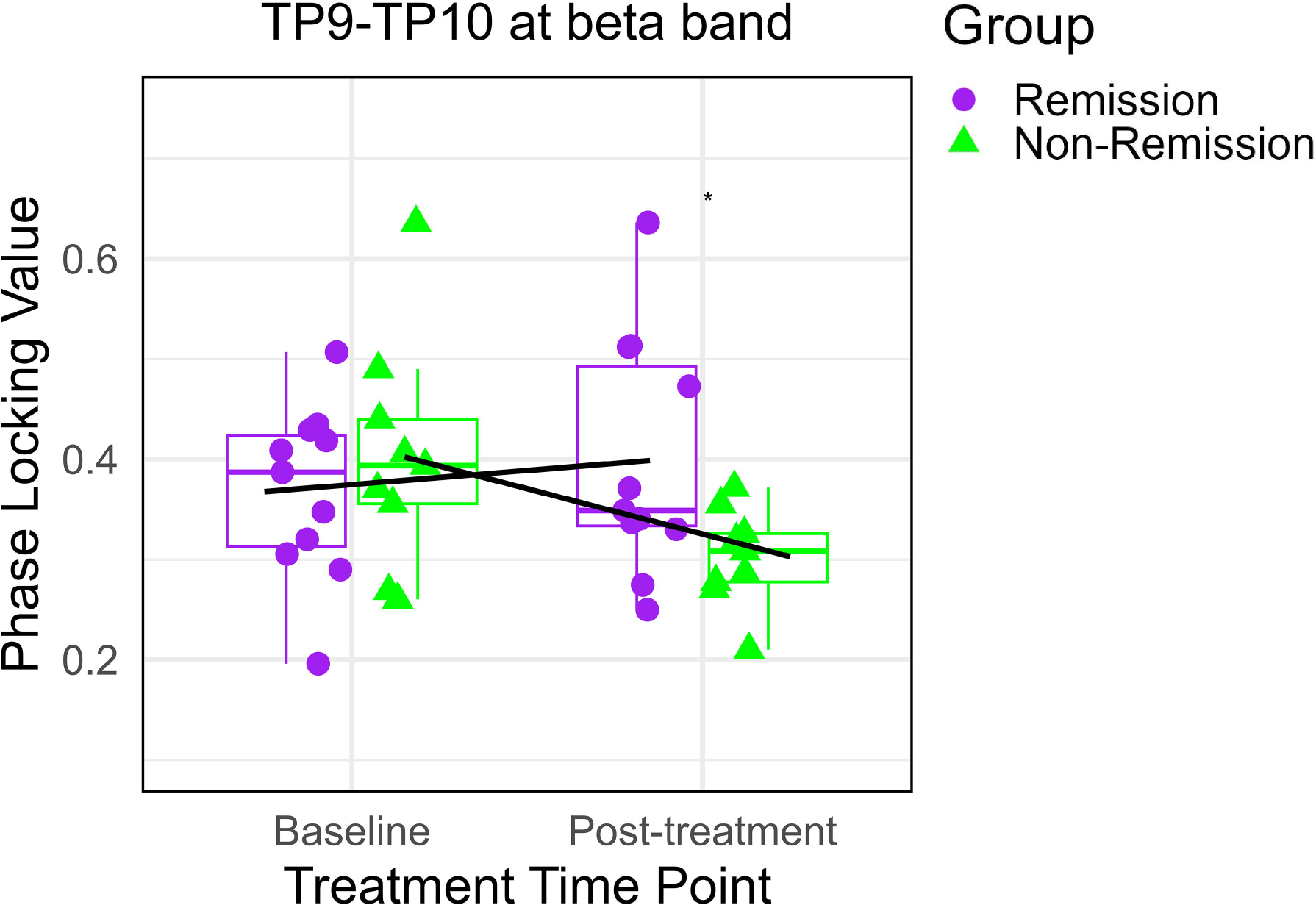
Comparative boxplot of phase locking value (PLV) of TP9 and TP10 at beta band between remission and non-remission groups.

### 3.6. Within group effects over time in remission and non-remission groups

In the remission group, a significant decrease in gamma band PLV AF8-TP9 was observed from baseline to post-treatment (*t* = 2.199, FDR-adjusted *p* = 0.044). Conversely, in the non-remission group, a significant decrease in beta band PLV TP9-TP10 was found from baseline to post-treatment (*t* = 2.397, FDR-adjusted *p* = 0.036) (Figure 2, Supplementary Table 3).

### 3.7. Deep learning-based prediction

The fully connected perceptron deep learning architecture, characterized by its fully connected layers, demonstrated superior performance metrics relative to the architecture predicated on a 1D-CNN. The fully connected perceptron architecture framework results are presented. Among the single EEG band PLV features, the highest classification accuracy of 62.2% was achieved for the alpha band (sensitivity 59.24%, specificity of 65.82%), and the beta band yielded the second highest classification accuracy of 59.93% (sensitivity of 68.94%, specificity 48.91%). In combinations of two band PLV, the theta-beta combination yielded the highest classification accuracy of 68.66% (sensitivity 73.85%, specificity 62.33%), and in the second highest accuracy was generated in the alpha-beta band feature combination accuracy 66.43% (sensitivity 74.52%, specificity 57.16%). Three band PLV combinations also exhibited high performance, particularly the combination of delta, alpha, and beta bands: accuracy 67.93% (sensitivity 74.49%, specificity 59.91%) and theta, alpha, and beta bands accuracy 67.25% (sensitivity 74.91%, specificity 57.88%). In four band PLV combination, theta, alpha, beta, gamma combination yielded the highest classification accuracy 67.29% (sensitivity 72.78%, specificity 60.58%) and the second highest classification accuracy 66.02% (sensitivity 73.37%, specificity 57.02%) for delta, alpha, beta, gamma. While combining PLV features from all the five classical EEG bands delta, theta, alpha, beta, and gamma, we obtained an accuracy 66.57 (sensitivity 68.24%, specificity 64.53%) (Supplementary Table 4).

## 4. Discussion

We investigated the relationship between EEG measures of brain activity and clinical outcomes to a home-based tDCS protocol in bipolar depression and whether baseline EEG measures could predictors of clinical remission to tDCS treatment. We examined EEG power and functional connectivity, as measured by PLV which quantifies phase interdependencies between brain regions.

We found a significant positive correlation with improvements in depressive symptoms and the delta band PLV in frontal and temporoparietal regional channel pairs. Furthermore, an interaction effect in network synchronisation was observed in beta band PLV in temporoparietal regions, in which participants who attained clinical remission showed increased synchronisation following tDCS treatment, which was decreased in participants who did not achieve clinical remission. The delta band is a low frequency EEG oscillation which is associated with cognitive control and enhanced internal concentration (Harmony et al., 2013), and beta band is a higher frequency oscillation which is associated with response preparation and inhibitory control, whereby increased inhibitory controls leads to increased beta band activity (Zhang et al., 2008; Tzagarakis et al., 2010). The findings suggest that that tDCS treatment enhances network synchronisation, potentially increasing inhibitory control, which underscores the improvement in depressive symptoms (Pellegrino et al., 2018).

Additional main effects of group were observed in several PLV frequency bands. In particular, participants who attained clinical remission following tDCS treatment showed increased PLV in channel pairs in frontal and temporal regions and in several frequency bands, including delta, theta, alpha and beta as compared to participants who did not achieve clinical remission following tDCS treatment. The present findings are consistent with reports of rTMS treatment in bipolar depression, in which PLV is increased in multiple channel pairs and in several frequency bands, namely theta, alpha, and beta bands, in participants who show a clinical response as compared to participants with who did not attain a clinical response (Zuchowicz et al., 2019). Increased theta and delta band activity in prefrontal and temporparietal regions has also been reported following rTMS treatment in bipolar depression in participants who showed a clinical response (Woźniak-Kwaśniewska et al., 2015). In contrast, participants who did not attain clinical remission following tDCS treatment showed a significant decrease in beta PLV, indicative of impaired neural phase synchronization, as compared to the participants who had attained clinical remission. Decreased synchronization in beta band was reported in bipolar disorder associated with cognitive impairments (Chen et al., 2008). The present findings indicate that there are underlying neural synchronizations in bipolar depression that can distinguish clinical outcome to tDCS.

Applying deep learning, we sought to explore whether we could predict clinical remission based on pre-treatment PLV features. Deep learning is a form of artificial intelligence that uses neural networks, which consists of a series of layers, to learn a representation of the data. The prediction accuracy ranged from 60-69%, while sensitivity values were generally higher, up to 76%. The highest prediction accuracy 69.45% was obtained for the combination of PLV features from the EEG bands theta, beta, and gamma with a balanced of sensitivity 71.68% and specificity of 66.72%. This finding aligns with another resting state EEG PLV analysis, in which the best classification performance for bipolar disorder was the beta band phase-synchronized feature (Duan et al., 2021). At the present time, we do not have any biomarkers that can help to identify and predict clinical responses. In unipolar depression, deep learning approaches have generated aggregate accuracies in the range of 70-80% for prediction of response to antidepressant medications, as measured by area under the curve (Squarcina et al., 2021). The findings suggest that adding a simple EEG measure at baseline can aid in identifying patients who will subsequently attain clinical remission following tDCS treatment, and in turn, this could identify patients who may not benefit from tDCS or who may require a combination of treatments.

The present study has several limitations. As the sample size was small and all participants had received active tDCS treatment, the power to detect a significant effect was limited and we cannot establish whether the findings are related to active or to placebo, sham effects. EEG data were acquired from four channels in a portable device which limited spatial resolution. We sought to include an easy-to-use device which participants could use at home that has strong reliability, research grade EEG data, and generates robust frequency measures comparable to a 64-channel device (Cannard et al., 2021; Krigolson et al., 2021). The EEG data were acquired in a resting state, while a cognitive task might have provided additional sensitivity in identifying predictors of clinical response (Mitoma et al., 2022).

In summary, tDCS treatment enhances network synchronisation, potentially increasing inhibitory control, which underscores improvement in depressive symptoms. Deep learning prediction models showed a range in which sensitivity values were generally higher, indicating that EEG-based measures might might aid predicting clinical response.

## Data Availability

All data generated in this study can be obtained by contacting the authors upon reasonable request.

