## Supplementary Materials for "Enhanced network synchronization connectivity following transcranial direct current stimulation (tDCS) in bipolar depression: effects on EEG oscillations and deep learning-based predictors of clinical remission"

**1. Methods**

**1.1 Aggregation of Resting State EEG Measures**

The eyes closed resting state EEG signals were utilized for the study to mitigate the potential artifacts caused from eye blinks. From the two five-minute eyes closed EEG recordings, measures including EEG band power and Phase Locking Value (PLV), a functional connectivity index, were extracted as EEG metrics. To compute these EEG measures, each EEG signal was segmented into 10-second windows, and following signal quality assessment, the EEG measures were computed. EEG band power was calculated for all four electrodes (AF7, AF8, TP9, TP10), while PLV was computed for all possible electrode pairs (AF7-AF8, AF7-TP9, AF7-TP10, AF8-TP9, AF8-TP10, TP9-TP10). These measures were extracted across all EEG bands, including the full band, δ, θ, α, 𝛽, and 𝛾, for subsequent analysis. Given the six EEG bands and four electrodes, a total of 24 band power values were calculated from a single raw EEG window. For PLV, considering six electrode pair combinations and six EEG bands, a total of 36 PLVs were computed. Subsequently, all EEG measures calculated from each EEG window were averaged across all windows per individual, resulting in 60 EEG measures for a 10-minute EEG recording. As both pre-treatment/baseline and post-treatment/week 6 EEG recordings spanned 10 minutes each, two sets of 60 EEG measures were available for statistical analysis. For the deep learning analysis, non-averaged EEG measures were utilized. Consequently, for each participant, there were 60 EEG windows extracted from a 10-minute EEG recording, prior to quality analysis. In an ideal scenario where all windows meet the quality criteria, each participant would contribute 60 * 60 EEG measures for deep learning training and testing.

1.2 **Band Power Value Calculation**

The EEG signal obtained from an electrode is a one-dimensional time series signal. We employ Equation (1) to compute the EEG band power value $P$ from the windowed and band-pass filtered EEG signals with a length of $N$:

$P_{x}= \frac{1}{N}\left( x^{T}. x \right)$	(1)

In Equation (1), $x$ represents the windowed and band-pass filtered EEG signal from an electrode, depicted as a column vector, while $x^{T}$ denotes its transpose. Subsequently, the power values are computed and averaged over all windows of each participant for a specific frequency band, resulting in the final power value corresponding to that band. Consequently, each participant will have six power values available, each corresponding to a distinct frequency band.

1.3 **PLV Calculation**

The phase locking value (PLV) assesses the phase synchrony between two time series signals (Hoke et al., 1989; Lachaux et al., 1999). It is a common measure employed to determine functional connectivity between EEG signals recorded from two electrodes, offering insight into the temporal relationships of neural signals independent of their amplitude. In this context, PLV is utilized to evaluate the frequency band-specific relationships between cortical regions, capturing the long-range synchronization of neural activity derived from EEG data. PLV is a statistical metric constrained within the range of 0 to 1. A PLV value approaching 1 indicates minimal variation in phase difference across the two EEG signals, while a value close to 0 suggests otherwise. The calculation of single trial PLV samples is as follows:

$PLV =\left| \frac{1}{N}\sum_{k=1}^{N} e^{j\Delta\theta(k)} \right|$	(2)

$\Delta\theta\left( k \right)=\theta_{1}\left( k \right)-\theta_{2}(k)$	(3)

In the above equations, $\theta_{1}\left( k \right)$ and $\theta_{2}(k)$ represent the instantaneous phase of two analytical signals derived from their respective signals. These analytical signals are computed from the real signals using the Hilbert transform (Boashash, 2003). PLV values are computed for each pair of electrodes, resulting in one PLV value calculated for every EEG window filtered in a specific EEG band. This process yields a total of six PLV values per EEG window. By averaging the PLV values over all windows, there will be two sets of 32 PLV values: one from the pre-treatment EEG data and the other from the week 6 EEG data for each participant. The average PLV values are used for statistical analysis and the non-averaged PLV values are used for deep learning analysis.

1.4 **Deep learning-based treatment prediction**

The study leveraged pre-treatment resting-state eyes closed EEG data from a cohort of 20 participants to classify treatment remission utilizing deep learning techniques. Among these participants, 11 achieved treatment remission following a 6-week course of tDCS treatment, while 9 did not exhibit remission. Treatment remission was operationally defined as having a MADRS score below 10 in week 6. For model training and testing, participants were categorized into two groups based on their remission status following tDCS treatment, as determined by their MADRS score at the conclusion of the 6-week trial. To assess the predictive utility of EEG-derived functional connectivity features, specifically Phase Locking Value (PLV), in treatment remission prognosis, PLV values were extracted from diverse EEG frequency bands of the pre-treatment resting-state EEG data. Two five-minute pre-treatment eyes closed resting-state EEG recordings were employed, constituting a 10-minute EEG signal segmented into 60 non-overlapping windows, each spanning 10 seconds. Prior to feature extraction, these EEG windows underwent screening using the Head Surface Index (HSI). Subsequently, PLV features extracted from various EEG bands, encompassing the full frequency band (1-60 Hz) and specific bands (delta, theta, alpha, beta, and gamma), were employed as inputs for the deep learning framework.

From each specific EEG band, PLV feature vectors with a dimensionality of 6 are generated, representing each of the six electrode pairs. These six-dimensional feature vectors, both individually and through the concatenation of PLV features from multiple EEG bands, were employed as inputs for deep learning models with varying parameters. This concatenation process at the feature level led to a linear increase in the feature dimension. To assess the effectiveness of different combinations of PLV features from individual bands and combinations of multiple bands, two distinct deep learning architectures were investigated. The first architecture utilized a fully connected perceptron deep learning structure, while the second employed a one-dimensional convolutional neural network (1DCNN) architecture. Considering the combination of features from multiple EEG bands, the dimensionality varied as follows: 6 (single band), 12 (two-band combination), 18 (three-band combination), 24 (four-band combination), and 30 (combination of all bands).

After conducting post-hoc analysis using paired t-tests, it was observed that specific PLV features extracted from the pre-treatment EEG exhibit statistically significant differences between the remission and non-remission groups within a 95% confidence interval. Consequently, PLV values from electrode pairs such as AF7-TP9 from the full band, AF7-AF8 from theta, AF7-TP10 from theta, AF7-TP10 from alpha, TP9-TP10 from alpha, and AF7-TP9 from beta were aggregated to form the feature vector. This concatenated feature set for each electrode pair had a dimensionality of six and was utilized to train the deep learning network for remission prediction in this study. The training process involved employing similar network parameters as those used for other six-dimensional features.

For the fully connected perceptron deep learning network, a four-layer architecture was implemented, comprising layers with 32, 32, 16, and 1 perceptrons (output layer) for all input combinations, except in the case of the all-band combination where the feature size was 3. In this scenario, 64 perceptrons were employed in the first layer. The all-band combination encompassed all PLV features extracted from the delta to gamma bands. In the 1DCNN-based architecture, the initial fully connected layer of the perceptron network was replaced with a convolutional layer. This convolutional layer employed a kernel size of 3. To ensure kernel overlap, the number of filters used was determined by multiplying the input dimension by a multiplication factor 2/3. Following the convolutional layer, a MaxPooling1D layer with a pool size of 2 was integrated to downsample the feature maps, aiming to extract the most relevant features while reducing computational complexity. For single-band PLV features with an input dimension of 6, 4 filters with a kernel size of 3 were used. Similarly, for dimensions 12, 18, 24, and 30, 8, 12, 16, and 20 filters were employed respectively. The activation function 'relu' was applied to all layers except the output layer, where 'sigmoid' was utilized. To minimize the 'binary cross-entropy' loss function, the 'adam' optimizer was employed.

**2. Statistical Analysis**

**2.1 Treatment Group Differences in Demographic and Clinical Characteristics**

To evaluate variances between the remission and non-remission groups regarding factors such as age, gender, MADRS, we performed statistical tests, two sample t test across the groups for continuous variables (e.g., age and MADRS overall score), chi-square tests for categorical variables (e.g., sex). The outcomes were presented in Table 1.

The Two Sample t-test was employed to assess the distinction in age between the remission and non-remission groups. The findings indicated a lack of significance (*t* = -1.90, *p* = .08), suggesting no notable difference in age between the treatment groups. The mean age estimate for the remission group was 47.19, compared to 55.11 for the non-remission group. Additionally, the Two Sample t-test was utilized to evaluate disparities in baseline MADRS scores between the remission and non-remission groups. However, there is a statistically different across treatment groups in baseline MADRS score (*t* = -2.44, *p* = .026) The baseline MADRS score for the remission group was 24.00, whereas for the non-remission group, it was 26.89.

The chi-squared test with Yates' continuity correction was employed to examine the association between sex and treatment group in the dataset. The analysis revealed a non-significant result (χ² = .62, *df* = 1, *p* = .43), indicating that there is no substantial association between sex and the assignment to treatment groups. This suggests that the distribution of gender is similar across both treatment groups.

**Supplementary Materials**

**Supplementary Table 1.**

Means, Standard Deviations, and main effect of Response Group (Remission/Non-Remission), main effect of time and interaction from Two-Way Analyses of Variance in Power and Phase Locking Value (PLV)

| Variable | Main effect of Group | | | | | | Main effect of Time | | | | | | Interaction between group and time | |
| --- | --- | --- | --- | --- | --- | --- | --- | --- | --- | --- | --- | --- | --- | --- |
|  | Baseline | | Post-Treatment | | *F (*1,1) | *p* | Baseline | | Post-treatment | | *F (*1,1) | *p* | *F (*1,1) | *p* |
|  | *M* | *SD* | *M* | *SD* |  |  | *M* | *SD* | *M* | *SD* |  |  |  |  |
| Full Power |  |  |  |  |  |  |  |  |  |  |  |  |  |  |
| AF7 | 979.59 | 1879.99 | 5896.89 | 13995.99 | 2.72 | 0.11 | 1744.76 | 4164.32 | 4640 | 13039.17 | 0.95 | 0.34 | 1.77 | 0.19 |
| AF8 | 656.01 | 992.82 | 4211.9 | 11250.31 | 2.08 | 0.16 | 2088.16 | 6674.95 | 2424.17 | 8735.55 | 0.02 | 0.89 | 0.11 | 0.74 |
| TP9 | 4163.87 | 6199.73 | 3383.5 | 11134.19 | 0.08 | 0.78 | 1622.24 | 2371.9 | 6003.17 | 11747.06 | 2.55 | 0.12 | 0.12 | 0.73 |
| TP10 | 3111.23 | 6711.04 | 3225.81 | 10177.27 | 0 | 0.97 | 1082.47 | 1487.63 | 5243.11 | 11451.45 | 2.46 | 0.13 | 0 | 0.95 |
| Delta Power |  |  |  |  |  |  |  |  |  |  |  |  |  |  |
| AF7 | 225.74 | 324.61 | 2514.76 | 6732.03 | 2.67 | 0.11 | 318.43 | 581.2 | 2193.17 | 6419.4 | 1.81 | 0.19 | 1.94 | 0.17 |
| AF8 | 202.16 | 365.73 | 651.81 | 1916.94 | 1.17 | 0.29 | 232.72 | 417.55 | 576.29 | 1817.84 | 0.69 | 0.41 | 1.56 | 0.22 |
| TP9 | 606.63 | 1089.53 | 806.81 | 2901.12 | 0.09 | 0.77 | 427.39 | 1142.9 | 966.03 | 2721.13 | 0.66 | 0.42 | 1.51 | 0.23 |
| TP10 | 288.12 | 237.04 | 800.76 | 2763.88 | 0.76 | 0.39 | 184.15 | 144.84 | 853.47 | 2603.3 | 1.31 | 0.26 | 1.13 | 0.3 |
| Theta Power |  |  |  |  |  |  |  |  |  |  |  |  |  |  |
| AF7 | 61.65 | 69.33 | 533.04 | 1688.77 | 1.76 | 0.19 | 68.25 | 116.62 | 479.3 | 1603.21 | 1.35 | 0.25 | 1.48 | 0.23 |
| AF8 | 43.14 | 41.28 | 644.9 | 2526.67 | 1.26 | 0.27 | 47.64 | 54.44 | 580.21 | 2397.99 | 1 | 0.32 | 1.29 | 0.26 |
| TP9 | 193.32 | 263.71 | 891.43 | 3553.51 | 0.86 | 0.36 | 120.56 | 240 | 894.38 | 3355.04 | 1.06 | 0.31 | 1.26 | 0.27 |
| TP10 | 94.83 | 78.48 | 747.11 | 2917.47 | 1.12 | 0.3 | 67.52 | 53.38 | 709.2 | 2761.27 | 1.09 | 0.3 | 1.26 | 0.27 |
| Alpha Power |  |  |  |  |  |  |  |  |  |  |  |  |  |  |
| AF7 | 42.59 | 57.25 | 335.06 | 1093.17 | 1.61 | 0.21 | 52.05 | 97.42 | 296.35 | 1037.56 | 1.13 | 0.29 | 1.59 | 0.22 |
| AF8 | 28.4 | 25.59 | 336.4 | 1311.03 | 1.23 | 0.27 | 26.74 | 26.86 | 307.26 | 1243.34 | 1.03 | 0.32 | 1.33 | 0.26 |
| TP9 | 78.73 | 78.69 | 310.82 | 1148.13 | 0.91 | 0.35 | 56.48 | 70.39 | 309.87 | 1084.25 | 1.09 | 0.3 | 1.28 | 0.26 |
| TP10 | 51.78 | 33.78 | 535.7 | 2098.52 | 1.19 | 0.28 | 42.08 | 26.26 | 497.01 | 1988.44 | 1.06 | 0.31 | 1.26 | 0.27 |
| Beta Power |  |  |  |  |  |  |  |  |  |  |  |  |  |  |
| AF7 | 258.56 | 700.86 | 702.3 | 1623.54 | 1.31 | 0.26 | 431.8 | 1057.62 | 484.69 | 1372.55 | 0.02 | 0.89 | 1.12 | 0.3 |
| AF8 | 118.87 | 125.84 | 332.4 | 1025.01 | 0.94 | 0.34 | 123.5 | 158.09 | 306.42 | 968.87 | 0.7 | 0.41 | 1.31 | 0.26 |
| TP9 | 150.93 | 112 | 306.58 | 1000.81 | 0.54 | 0.47 | 106.97 | 109.2 | 334.98 | 939.89 | 1.16 | 0.29 | 1.42 | 0.24 |
| TP10 | 123.19 | 101.42 | 288.41 | 923.51 | 0.71 | 0.41 | 95.97 | 92.99 | 299.11 | 870.81 | 1.08 | 0.31 | 1.52 | 0.23 |
| Gamma Power |  |  |  |  |  |  |  |  |  |  |  |  |  |  |
| AF7 | 401.3 | 1094.32 | 2191.19 | 6506.23 | 1.57 | 0.22 | 938.12 | 2613.45 | 1475.38 | 5821.17 | 0.14 | 0.71 | 0.65 | 0.42 |
| AF8 | 282.12 | 572.64 | 2335.89 | 7816.87 | 1.49 | 0.23 | 1891.79 | 7302.42 | 520.84 | 1707.94 | 0.67 | 0.42 | 0.55 | 0.46 |
| TP9 | 3531.54 | 6885.68 | 881.86 | 1771.02 | 2.65 | 0.11 | 986.15 | 1968.57 | 3692.22 | 7151.13 | 2.79 | 0.1 | 1.21 | 0.28 |
| TP10 | 2909.37 | 7410.89 | 711.58 | 1714.81 | 1.58 | 0.22 | 765.94 | 1663.66 | 3074.78 | 7763.82 | 1.76 | 0.19 | 1.96 | 0.17 |
| Full PLV |  |  |  |  |  |  |  |  |  |  |  |  |  |  |
| AF7-AF8 | 0.289 | 0.186 | 0.157 | 0.194 | 3.45 | 0.071 | 0.283 | 0.202 | 0.227 | 0.107 | 2.194 | 0.147 | 0.18 | 0.674 |
| AF7-TP9 | **0.196** | **0.293** | **0.102** | **0.158** | **5.762*** | **0.022** | 0.208 | 0.271 | 0.12 | 0.149 | 2.491 | 0.123 | 1.816 | 0.186 |
| AF7-TP10 | 0.168 | 0.208 | 0.084 | 0.166 | .968 | 0.332 | 0.181 | 0.191 | 0.116 | 0.141 | 0.062 | 0.804 | 1.004 | 0.323 |
| AF8-TP9 | 0.169 | 0.175 | 0.138 | 0.143 | .022 | 0.883 | 0.14 | 0.204 | 0.093 | 0.169 | 2.102 | 0.156 | 0 | 0.987 |
| AF8-TP10 | 0.238 | 0.242 | 0.127 | 0.109 | .016 | 0.902 | 0.226 | 0.254 | 0.102 | 0.132 | 0.522 | 0.475 | 0.04 | 0.843 |
| TP9-TP10 | 0.596 | 0.522 | 0.253 | 0.218 | .942 | 0.338 | 0.531 | 0.594 | 0.24 | 0.238 | 0.687 | 0.413 | 0.127 | 0.723 |
| Delta PLV |  |  |  |  |  |  |  |  |  |  |  |  |  |  |
| AF7-AF8 | **0.385** | **0.264** | **0.152** | **0.09** | **8.874**** | **0.005** | 0.342 | 0.319 | 0.172 | 0.101 | 0.328 | 0.571 | 2.069 | 0.159 |
| AF7-TP9 | 0.353 | 0.306 | 0.134 | 0.043 | 2.015 | 0.164 | 0.338 | 0.325 | 0.125 | 0.084 | 0.141 | 0.71 | 1.775 | 0.191 |
| AF7-TP10 | **0.345** | **0.245** | **0.15** | **0.057** | **6.939*** | **0.012** | 0.317 | 0.284 | 0.15 | 0.1 | 0.762 | 0.388 | 0.733 | 0.398 |
| AF8-TP9 | **0.311** | **0.255** | **0.098** | **0.063** | **4.346*** | **0.044** | 0.287 | 0.285 | 0.103 | 0.073 | 0.009 | 0.926 | 1.644 | 0.208 |
| AF8-TP10 | 0.344 | 0.318 | 0.109 | 0.073 | 0.725 | 0.4 | 0.338 | 0.326 | 0.111 | 0.076 | 0.145 | 0.706 | 0.511 | 0.479 |
| TP9-TP10 | **0.608** | **0.49** | **0.155** | **0.158** | **5.62*** | **0.023** | 0.57 | 0.54 | 0.179 | 0.154 | 0.347 | 0.559 | 1.483 | 0.231 |
| Theta PLV |  |  |  |  |  |  |  |  |  |  |  |  |  |  |
| AF7-AF8 | **0.323** | **0.185** | **0.16** | **0.057** | **11.917**** | **0.001** | 0.276 | 0.245 | 0.168 | 0.112 | 0.603 | 0.443 | 0.738 | 0.396 |
| AF7-TP9 | 0.339 | 0.29 | 0.143 | 0.067 | 1.711 | 0.199 | 0.318 | 0.316 | 0.129 | 0.107 | 0.001 | 0.973 | 0.577 | 0.453 |
| AF7-TP10 | 0.258 | 0.154 | 0.141 | 0.034 | 9.001 | 0.005 | 0.216 | 0.207 | 0.141 | 0.095 | 0.074 | 0.787 | 0.521 | 0.475 |
| AF8-TP9 | 0.235 | 0.191 | 0.102 | 0.075 | 2.217 | 0.145 | 0.21 | 0.22 | 0.092 | 0.094 | 0.104 | 0.749 | 0.821 | 0.371 |
| AF8-TP10 | 0.322 | 0.319 | 0.122 | 0.069 | 0.006 | 0.939 | 0.32 | 0.322 | 0.108 | 0.095 | 0.004 | 0.953 | 0 | 0.989 |
| TP9-TP10 | **0.551** | **0.42** | **0.143** | **0.146** | **8.12*** | **0.007** | 0.506 | 0.479 | 0.172 | 0.144 | 0.34 | 0.563 | 1.59 | 0.215 |
| Alpha PLV |  |  |  |  |  |  |  |  |  |  |  |  |  |  |
| AF7-AF8 | **0.312** | **0.177** | **0.168** | **0.073** | **9.777**** | **0.003** | 0.264 | 0.239 | 0.163 | 0.136 | 0.345 | 0.561 | 0.726 | 0.4 |
| AF7-TP9 | **0.39** | **0.308** | **0.129** | **0.078** | **5.393*** | **0.026** | 0.343 | 0.363 | 0.141 | 0.084 | 0.329 | 0.57 | 0.444 | 0.509 |
| AF7-TP10 | **0.251** | **0.162** | **0.121** | **0.039** | **8.515**** | **0.006** | 0.21 | 0.211 | 0.122 | 0.082 | 0 | 0.984 | 0.491 | 0.488 |
| AF8-TP9 | 0.246 | 0.19 | 0.112 | 0.058 | 3.486 | 0.07 | 0.219 | 0.224 | 0.103 | 0.089 | 0.03 | 0.863 | 0.315 | 0.578 |
| AF8-TP10 | 0.378 | 0.337 | 0.11 | 0.09 | 1.533 | 0.224 | 0.37 | 0.349 | 0.104 | 0.102 | 0.395 | 0.534 | 0.016 | 0.901 |
| TP9-TP10 | **0.527** | **0.42** | **0.135** | **0.088** | **8.032**** | **0.007** | 0.49 | 0.468 | 0.14 | 0.114 | 0.337 | 0.565 | 0.053 | 0.818 |
| Beta PLV |  |  |  |  |  |  |  |  |  |  |  |  |  |  |
| AF7-AF8 | 0.291 | 0.206 | 0.154 | 0.167 | 2.772 | 0.105 | 0.275 | 0.231 | 0.193 | 0.129 | 0.752 | 0.392 | 0.331 | 0.569 |
| AF7-TP9 | **0.341** | **0.237** | **0.108** | **0.115** | **8.322**** | **0.007** | 0.295 | 0.293 | 0.136 | 0.109 | 0.005 | 0.944 | 0.809 | 0.374 |
| AF7-TP10 | **0.255** | **0.166** | **0.102** | **0.105** | **7.235*** | **0.011** | 0.227 | 0.203 | 0.121 | 0.102 | 0.549 | 0.464 | 1.025 | 0.318 |
| AF8-TP9 | 0.222 | 0.195 | 0.109 | 0.095 | 0.733 | 0.397 | 0.207 | 0.213 | 0.108 | 0.1 | 0.034 | 0.855 | 2.335 | 0.135 |
| AF8-TP10 | 0.32 | 0.282 | 0.089 | 0.098 | 1.685 | 0.203 | 0.31 | 0.295 | 0.083 | 0.106 | 0.269 | 0.607 | 2.447 | 0.126 |
| TP9-TP10 | 0.383 | 0.353 | 0.102 | 0.099 | 0.991 | 0.326 | 0.356 | 0.383 | 0.104 | 0.099 | 0.813 | 0.373 | **4.476*** | **0.041** |
| Gamma PLV |  |  |  |  |  |  |  |  |  |  |  |  |  |  |
| AF7-AF8 | 0.336 | 0.265 | 0.172 | 0.209 | 1.337 | 0.255 | 0.321 | 0.286 | 0.233 | 0.14 | 0.321 | 0.575 | 0.027 | 0.872 |
| AF7-TP9 | 0.271 | 0.389 | 0.16 | 0.286 | 2.734 | 0.107 | 0.279 | 0.369 | 0.204 | 0.251 | 1.608 | 0.213 | 0.075 | 0.785 |
| AF7-TP10 | 0.244 | 0.361 | 0.167 | 0.273 | 2.678 | 0.11 | 0.275 | 0.318 | 0.193 | 0.259 | 0.359 | 0.553 | 0.247 | 0.622 |
| AF8-TP9 | 0.294 | 0.277 | 0.218 | 0.209 | 0.07 | 0.792 | **0.207** | **0.366** | **0.161** | **0.23** | **6.138*** | **0.018** | 0.255 | 0.617 |
| AF8-TP10 | 0.332 | 0.291 | 0.232 | 0.165 | 0.386 | 0.539 | 0.274 | 0.353 | 0.166 | 0.232 | 1.478 | 0.232 | 0.028 | 0.868 |
| TP9-TP10 | 0.631 | 0.605 | 0.349 | 0.271 | 0.071 | 0.791 | **0.514** | **0.724** | **0.32** | **0.274** | **4.766*** | **0.036** | 0.438 | 0.512 |

*Note. ^*^p < .05, ^**^p < .01, p values are corrected for False Discovery Rate (FDR)*

**Supplementary Materials**

**Supplementary Table 2.**

Results of post-hoc test Examining the Treatment group difference at the Baseline and Post-treatment for variables with significant main effect of group in two-way Anova

| PLV Variables | Baseline | | | | *t* | *p* | Post-Treatment | | | | *t* | *p* |
| --- | --- | --- | --- | --- | --- | --- | --- | --- | --- | --- | --- | --- |
|  | Remission | | Non-Remission | |  |  | Remission | | Non-Remission | |  |  |
|  | *M* | *SD* | *M* | *SD* |  |  | *M* | *SD* | *M* | *SD* |  |  |
| Full |  |  |  |  |  |  |  |  |  |  |  |  |
| AF7-TP9 | **0.20** | **0.12** | **0.35** | **0.15** | **-2.49*** | **0.02** | 0.19 | 0.09 | 0.23 | 0.15 | -0.74 | 0.47 |
| Delta |  |  |  |  |  |  |  |  |  |  |  |  |
| AF7-AF8 | 0.35 | 0.09 | 0.29 | 0.11 | 1.37 | 0.19 | 0.42 | 0.19 | 0.24 | 0.06 | 2.88* | **0.01** |
| AF7-TP10 | 0.31 | 0.11 | 0.25 | 0.07 | 1.62 | 0.12 | 0.38 | 0.18 | 0.24 | 0.04 | 2.34* | **0.04** |
| AF8-TP9 | 0.29 | 0.07 | 0.27 | 0.08 | 0.64 | 0.53 | 0.33 | 0.12 | 0.24 | 0.04 | 2.32* | **0.04** |
| TP9-TP10 | 0.57 | 0.15 | 0.51 | 0.17 | 0.81 | 0.43 | 0.65 | 0.16 | 0.47 | 0.16 | 2.51* | **0.02** |
| Theta |  |  |  |  |  |  |  |  |  |  |  |  |
| AF7-AF8 | **0.29** | **0.13** | **0.18** | **0.06** | **2.44*** | **0.03** | **0.35** | **0.19** | **0.18** | **0.06** | **2.86*** | **0.01** |
| AF7-TP10 | **0.24** | **0.11** | **0.16** | **0.04** | **2.16*** | **0.05** | **0.27** | **0.17** | **0.15** | **0.02** | **2.51*** | **0.03** |
| TP9-TP10 | 0.51 | 0.17 | 0.44 | 0.16 | 1.10 | 0.29 | **0.59** | **0.15** | **0.40** | **0.13** | **2.93*** | **0.01** |
| Alpha |  |  |  |  |  |  |  |  |  |  |  |  |
| AF7-AF8 | 0.28 | 0.16 | 0.19 | 0.08 | 1.78 | 0.09 | 0.34 | 0.18 | 0.17 | 0.07 | 2.93* | **0.01** |
| AF7-TP9 | 0.39 | 0.10 | 0.33 | 0.05 | 1.71 | 0.11 | 0.39 | 0.16 | 0.29 | 0.10 | 1.83 | 0.08 |
| AF7-TP10 | 0.24 | 0.10 | 0.17 | 0.04 | 2.11 | **0.05** | 0.26 | 0.15 | 0.15 | 0.03 | 2.41* | **0.03** |
| TP9-TP10 | **0.51** | **0.12** | **0.41** | **0.08** | **2.15*** | **0.04** | 0.54 | 0.15 | 0.43 | 0.10 | 2.06 | 0.05 |
| Beta |  |  |  |  |  |  |  |  |  |  |  |  |
| AF7-TP9 | **0.33** | **0.09** | **0.25** | **0.12** | **2.54*** | **0.02** | 0.36 | 0.13 | 0.22 | 0.11 | 1.17 | 0.26 |
| AF7-TP10 | 0.23 | 0.08 | 0.17 | 0.12 | 1.17 | 0.26 | 0.28 | 0.12 | 0.16 | 0.09 | 2.60* | **0.02** |

*Note. ^*^p < .05, ^**^p < .01, p values are adjusted by FDR*

Post-hoc analyses were conducted for each significant variable identified, and the outcomes are presented in Supplementary Table 2. Specifically, we previously found a significant main effect for group in the PLV of AF7-TP9 across the full, alpha, and beta bands. Based on t-tests conducted across groups at both baseline and post-treatment, we observed that the significant group effect stems from differences between groups at baseline (*t_full_* = -2.49, FDR-adjusted *p* = .02; *t_beta_* = 2.54, FDR-adjusted *p* = .02), as there were no significant differences post-treatment. Similarly, variables exhibiting significant group differences at baseline, but not post-treatment include: PLV of TP9-TP10 in the alpha band (*t* = 2.15, FDR-adjusted *p* = .04). Moreover, PLV of AF7-AF8 and AF7-TP10 in theta band show both significant group differences at baseline (*t_AF7-AF8_* = 2.44, FDR-adjusted *p* = .03; *t_AF7-TP10_* = 2.16, FDR-adjusted *p* = .05) and post-treatment (*t_AF7-AF8_* = 2.86, FDR-adjusted *p* = .01; *t_AF7 -TP10_* = 2.51, FDR-adjusted *p* = .03).

Importantly, we focus on the variables that show significant group differences at post-treatment, but not at baseline. Specifically, BP remission group have a generally greater PLV value compared to the BP non-remission group at the post-treatment, including PLV of AF7-AF8 in both delta and alpha bands (*t_delta_* = 2.88, FDR-adjusted *p* = .01; *t_alpha_* = 2.93, FDR-adjusted *p* = .01); PLV of AF7-TP10 in delta and beta bands (*t_delta_* = 2.34, FDR-adjusted *p* = .04; *t_beta_* = 2.60, FDR-adjusted *p* = .02); PLV of TP9-TP10 in delta and theta bands (*t_delta_* = 2.51, FDR-adjusted *p* = .02; *t_theta_* = 2.93, FDR-adjusted *p* = .01) and PLV of AF8-TP9 in the delta band (*t* = 2.32, FDR-adjusted *p* = .04).

**Supplementary Materials**

**Supplementary Table 3.**

Change in PLV from the baseline to post-treatment within each remission group from paired two-sample t test

| Variable | Remission Group | | | | | | Non - Remission Group | | | | | |
| --- | --- | --- | --- | --- | --- | --- | --- | --- | --- | --- | --- | --- |
|  | Baseline | | Post-Treatment | | *t* | *p* | Baseline | | Post-Treatment | | *t* | *p* |
|  | *M* | *SD* | *M* | *SD* |  |  | *M* | *SD* | *M* | *SD* |  |  |
| Full  Power |  |  |  |  |  |  |  |  |  |  |  |  |
| AF7 | 645.11 | 677.61 | 1314.08 | 2591.69 | -0.83 | 0.42 | 9522.64 | 18827.09 | 2271.14 | 5676.56 | 1.11 | 0.30 |
| AF8 | 454.35 | 345.10 | 857.68 | 1364.33 | -0.95 | 0.36 | 4831.73 | 13008.87 | 3592.07 | 9943.09 | 0.23 | 0.82 |
| TP9 | 5924.08 | 8082.32 | 2403.66 | 2928.61 | 1.36 | 0.20 | 6099.83 | 15686.31 | 667.18 | 88.28 | 1.04 | 0.33 |
| TP10 | 5118.28 | 92.93 | 1104.18 | 1029.91 | 1.44 | 0.18 | 5395.68 | 14338.00 | 1055.93 | 1982.09 | 0.90 | 0.39 |
| Delta Power |  |  |  |  |  |  |  |  |  |  |  |  |
| AF7 | 285.38 | 431.25 | 166.10 | 165.77 | 0.86 | 0.41 | 9522.64 | 18827.09 | 2271.14 | 5676.56 | 1.11 | 0.30 |
| AF8 | 14.25 | 122.26 | 264.08 | 507.45 | -0.79 | 0.45 | 4831.73 | 13008.87 | 3592.07 | 9943.09 | 0.23 | 0.82 |
| TP9 | 507.74 | 425.71 | 705.52 | 1513.31 | -0.42 | 0.68 | 6099.83 | 15686.31 | 667.18 | 88.28 | 1.04 | 0.33 |
| TP10 | 342.62 | 294.80 | 233.62 | 156.69 | 1.08 | 0.30 | 5395.68 | 14338.00 | 1055.93 | 1982.09 | 0.90 | 0.39 |
| Theta Power |  |  |  |  |  |  |  |  |  |  |  |  |
| AF7 | 72.87 | 90.13 | 50.43 | 41.17 | 0.75 | 0.47 | 976.06 | 2364.22 | 90.03 | 170.91 | 1.12 | 0.29 |
| AF8 | 36.29 | 28.36 | 49.99 | 51.68 | -0.77 | 0.45 | 1245.01 | 3571.01 | 44.78 | 60.70 | 1.01 | 0.34 |
| TP9 | 198.75 | 219.36 | 187.88 | 312.82 | 0.09 | 0.93 | 1744.58 | 5019.46 | 38.28 | 25.08 | 1.02 | 0.34 |
| TP10 | 104.23 | 93.87 | 85.43 | 62.66 | 0.55 | 0.59 | 1448.61 | 4120.58 | 45.62 | 29.66 | 1.02 | 0.34 |
| Alpha Power |  |  |  |  |  |  |  |  |  |  |  |  |
| AF7 | 33.84 | 33.05 | 51.34 | 74.98 | -0.71 | 0.49 | 617.20 | 1531.28 | 52.93 | 124.55 | 1.10 | 0.30 |
| AF8 | 24.91 | 21.99 | 31.88 | 29.41 | -0.63 | 0.54 | 652.35 | 1851.29 | 2.46 | 23.48 | 1.02 | 0.34 |
| TP9 | 81.31 | 7.97 | 76.15 | 89.18 | 0.15 | 0.88 | 589.21 | 162.54 | 32.43 | 25.35 | 1.03 | 0.33 |
| TP10 | 55.07 | 4.86 | 48.50 | 26.50 | 0.45 | 0.66 | 1037.16 | 2965.07 | 34.25 | 25.17 | 1.01 | 0.34 |
| Beta Power |  |  |  |  |  |  |  |  |  |  |  |  |
| AF7 | 1.48 | 93.40 | 416.65 | 983.78 | -1.06 | 0.31 | 954.28 | 2003.08 | 45.31 | 1202.53 | 0.65 | 0.53 |
| AF8 | 96.89 | 92.27 | 14.85 | 153.88 | -0.81 | 0.43 | 562.51 | 1443.83 | 102.29 | 169.83 | 0.95 | 0.37 |
| TP9 | 15.66 | 106.43 | 151.20 | 122.54 | -0.01 | 0.99 | 56.26 | 1407.16 | 52.90 | 59.76 | 1.08 | 0.31 |
| TP10 | 115.99 | 101.37 | 13.40 | 105.88 | -0.33 | 0.75 | 522.93 | 1298.34 | 53.89 | 53.87 | 1.08 | 0.31 |
| Gamma Power |  |  |  |  |  |  |  |  |  |  |  |  |
| AF7 | 149.25 | 151.08 | 653.35 | 1533.70 | -1.08 | 0.30 | 3096.20 | 8664.72 | 1286.17 | 361.18 | 0.58 | 0.57 |
| AF8 | 159.79 | 151.26 | 404.45 | 795.51 | -1.00 | 0.34 | 962.12 | 2549.73 | 3709.65 | 10913.17 | -0.74 | 0.48 |
| TP9 | 5688.75 | 9108.56 | 1374.33 | 2522.45 | 1.51 | 0.16 | 1252.00 | 2359.43 | 511.72 | 888.76 | 0.88 | 0.40 |
| TP10 | 5166.93 | 10155.72 | 651.80 | 991.72 | 1.47 | 0.17 | 517.69 | 927.31 | 905.46 | 2303.09 | -0.47 | 0.65 |
| Full PLV |  |  |  |  |  |  |  |  |  |  |  |  |
| AF7-AF8 | 0.237 | 0.1 | 0.34 | 0.19 | -1.58 | 0.134 | 0.158 | 0.104 | 0.237 | -0.596 | 0.564 | 0.237 |
| AF7-TP9 | 0.203 | 0.118 | 0.189 | 0.088 | .323 | 0.75 | 0.354 | 0.146 | 0.203 | 1.741 | 0.101 | 0.203 |
| AF7-TP10 | 0.155 | 0.098 | 0.181 | 0.07 | -.738 | 0.47 | 0.236 | 0.176 | 0.155 | 0.697 | 0.496 | 0.155 |
| AF8-TP9 | 0.201 | 0.173 | 0.137 | 0.088 | 1.09 | 0.295 | 0.208 | 0.175 | 0.201 | 0.961 | 0.354 | 0.201 |
| AF8-TP10 | 0.255 | 0.146 | 0.22 | 0.108 | .632 | 0.535 | 0.252 | 0.121 | 0.255 | 0.366 | 0.719 | 0.255 |
| TP9-TP10 | 0.616 | 0.283 | 0.577 | 0.231 | .351 | 0.729 | 0.568 | 0.182 | 0.616 | 0.905 | 0.38 | 0.616 |
| Delta PLV |  |  |  |  |  |  |  |  |  |  |  |  |
| AF7-AF8 | 0.347 | 0.09 | 0.422 | 0.193 | -1.17 | 0.26 | 0.285 | 0.109 | 0.347 | 0.968 | 0.351 | 0.347 |
| AF7-TP9 | 0.327 | 0.108 | 0.379 | 0.158 | -.904 | 0.378 | 0.324 | 0.045 | 0.327 | 1.896 | 0.077 | 0.327 |
| AF7-TP10 | 0.314 | 0.112 | 0.376 | 0.181 | -.965 | 0.348 | 0.247 | 0.073 | 0.314 | 0.099 | 0.923 | 0.314 |
| AF8-TP9 | 0.294 | 0.073 | 0.328 | 0.12 | -.795 | 0.438 | 0.273 | 0.077 | 0.294 | 1.205 | 0.25 | 0.294 |
| AF8-TP10 | 0.328 | 0.079 | 0.359 | 0.134 | -.664 | 0.516 | 0.324 | 0.077 | 0.328 | 0.353 | 0.729 | 0.328 |
| TP9-TP10 | 0.566 | 0.146 | 0.65 | 0.16 | -1.285 | 0.214 | 0.509 | 0.166 | 0.566 | 0.493 | 0.628 | 0.566 |
| Theta PLV |  |  |  |  |  |  |  |  |  |  |  |  |
| AF7-AF8 | 0.292 | 0.126 | 0.354 | 0.19 | -.903 | 0.379 | 0.188 | 0.058 | 0.292 | 0.252 | 0.804 | 0.292 |
| AF7-TP9 | 0.326 | 0.134 | 0.353 | 0.158 | -.431 | 0.671 | 0.305 | 0.068 | 0.326 | 0.944 | 0.359 | 0.326 |
| AF7-TP10 | 0.242 | 0.111 | 0.274 | 0.17 | -.523 | 0.608 | 0.163 | 0.045 | 0.242 | 1.151 | 0.277 | 0.242 |
| AF8-TP9 | 0.227 | 0.101 | 0.242 | 0.107 | -.326 | 0.748 | 0.21 | 0.09 | 0.227 | 1.1 | 0.291 | 0.227 |
| AF8-TP10 | 0.323 | 0.116 | 0.321 | 0.133 | .045 | 0.965 | 0.32 | 0.068 | 0.323 | 0.045 | 0.965 | 0.323 |
| TP9-TP10 | 0.512 | 0.126 | 0.591 | 0.154 | -1.311 | 0.205 | 0.439 | 0.162 | 0.512 | 0.528 | 0.605 | 0.512 |
| Alpha PLV |  |  |  |  |  |  |  |  |  |  |  |  |
| AF7-AF8 | 0.283 | 0.159 | 0.341 | 0.179 | -.808 | 0.429 | 0.185 | 0.081 | 0.283 | 0.43 | 0.673 | 0.283 |
| AF7-TP9 | 0.389 | 0.098 | 0.39 | 0.159 | -.018 | 0.986 | 0.331 | 0.051 | 0.389 | 1.276 | 0.226 | 0.389 |
| AF7-TP10 | 0.241 | 0.095 | 0.26 | 0.147 | -.351 | 0.73 | 0.174 | 0.042 | 0.241 | 1.345 | 0.199 | 0.241 |
| AF8-TP9 | 0.241 | 0.104 | 0.251 | 0.124 | -.203 | 0.841 | 0.202 | 0.065 | 0.241 | 0.85 | 0.409 | 0.241 |
| AF8-TP10 | 0.366 | 0.103 | 0.39 | 0.12 | -.509 | 0.616 | 0.329 | 0.103 | 0.366 | -0.369 | 0.717 | 0.366 |
| TP9-TP10 | 0.512 | 0.12 | 0.542 | 0.152 | -.507 | 0.618 | 0.414 | 0.083 | 0.512 | -0.285 | 0.779 | 0.512 |
| Beta PLV |  |  |  |  |  |  |  |  |  |  |  |  |
| AF7-AF8 | 0.256 | 0.128 | 0.327 | 0.175 | -1.088 | 0.291 | 0.2 | 0.13 | 0.256 | -0.146 | 0.886 | 0.256 |
| AF7-TP9 | 0.325 | 0.091 | 0.357 | 0.125 | -.678 | 0.506 | 0.254 | 0.121 | 0.325 | 0.596 | 0.559 | 0.325 |
| AF7-TP10 | 0.228 | 0.08 | 0.282 | 0.117 | -1.272 | 0.22 | 0.172 | 0.121 | 0.228 | 0.244 | 0.81 | 0.228 |
| AF8-TP9 | 0.203 | 0.082 | 0.242 | 0.131 | -.831 | 0.417 | 0.225 | 0.122 | 0.203 | 1.385 | 0.195 | 0.203 |
| AF8-TP10 | 0.291 | 0.08 | 0.348 | 0.093 | -1.529 | 0.142 | 0.299 | 0.136 | 0.291 | 0.758 | 0.468 | 0.291 |
| TP9-TP10 | 0.368 | 0.086 | 0.399 | 0.119 | -.703 | 0.491 | **0.402** | **0.115** | 0.368 | **2.397*** | **0.036** | 0.368 |
| Gamma PLV |  |  |  |  |  |  |  |  |  |  |  |  |
| AF7-AF8 | 0.323 | 0.148 | 0.349 | 0.2 | -.342 | 0.736 | 0.242 | 0.123 | 0.323 | -0.454 | 0.659 | 0.323 |
| AF7-TP9 | 0.307 | 0.184 | 0.234 | 0.132 | 1.069 | 0.299 | 0.446 | 0.309 | 0.307 | 0.827 | 0.421 | 0.307 |
| AF7-TP10 | 0.249 | 0.204 | 0.239 | 0.131 | .145 | 0.886 | 0.402 | 0.304 | 0.249 | 0.623 | 0.542 | 0.249 |
| AF8-TP9 | **0.389** | **0.254** | **0.2** | **0.127** | **2.199*** | **0.044** | 0.339 | 0.207 | **0.389** | 1.271 | 0.222 | **0.389** |
| AF8-TP10 | 0.366 | 0.276 | 0.297 | 0.186 | .691 | 0.498 | 0.337 | 0.179 | 0.366 | 1.193 | 0.251 | 0.366 |
| TP9-TP10 | 0.764 | 0.301 | 0.497 | 0.355 | 1.907 | 0.071 | 0.675 | 0.247 | 0.764 | 1.098 | 0.289 | 0.764 |

*Note.* ^*^*p* < .05, ^**^*p* < .01, *p values are corrected for False Discovery Rate (FDR)*

**Supplementary Materials**

**Supplementary Table 4**

Deep learning classification results for the fully connected perceptron model

| EEG bands whose PLV  features are combined | Accuracy | Sensitivity | Specificity |
| --- | --- | --- | --- |
| Full band | 54.40 | 52.44 | 56.79 |
| Delta | 50.91 | 63.18 | 35.92 |
| Theta | 58.51 | 59.83 | 56.89 |
| Alpha | 62.20 | 59.24 | 65.82 |
| Beta | 59.93 | 68.94 | 48.91 |
| Gamma | 55.25 | 47.12 | 65.18 |
| Delta, Theta | 57.03 | 60.44 | 52.87 |
| Delta, Alpha | 56.07 | 62.51 | 48.68 |
| Delta, Beta | 63.45 | 72.61 | 52.25 |
| Delta, Gamma | 52.44 | 48.02 | 57.86 |
| Theta, Alpha | 61.04 | 60.61 | 61.57 |
| Theta, Beta | 68.66 | 73.85 | 62.33 |
| Theta, Gamma | 55.42 | 54.47 | 56.57 |
| Alpha, Beta | 66.43 | 74.02 | 57.16 |
| Alpha, Gamma | 51.91 | 46.38 | 58.67 |
| Beta, Gamma | 64.55 | 67.39 | 61.09 |
| Delta, Theta, Alpha | 59.07 | 61.50 | 56.10 |
| Delta, Theta, Beta | 62.23 | 68.97 | 54.00 |
| Delta, Theta, Gamma | 52.92 | 52.86 | 52.99 |
| Delta, Alpha, Beta | 67.93 | 74.49 | 59.91 |
| Delta, Alpha, Gamma | 53.51 | 51.20 | 56.35 |
| Delta, Beta, Gamma | 60.99 | 61.10 | 60.86 |
| Theta, Alpha, Beta | 67.25 | 74.91 | 57.88 |
| Theta, Alpha, Gamma | 51.64 | 48.00 | 56.08 |
| Theta, Beta, Gamma | 69.45 | 71.68 | 66.72 |
| Alpha, Beta, Gamma | 63.06 | 66.81 | 58.47 |
| Delta, Theta, Alpha, Beta | 61.68 | 67.58 | 54.48 |
| Delta, Theta, Alpha, Gamma | 52.56 | 48.60 | 57.40 |
| Delta, Theta, Beta, Gamma | 63.47 | 69.76 | 55.79 |
| Delta, Alpha, Beta, Gamma | 66.02 | 73.37 | 57.02 |
| Theta, Alpha, Beta, Gamma | 67.29 | 72.78 | 60.58 |
| Delta, Theta, Alpha, Beta, Gamma | 66.57 | 68.24 | 64.53 |

**Supplementary Materials**

**Supplementary Table 5.**

Pearson’s Correlation between change in EEG metrics and proportional change in MADRS score across all subjects

| Variable | Correlation Coefficient | *p* |
| --- | --- | --- |
| Full Power |  |  |
| AF7 | 0.09 | 0.71 |
| AF8 | -0.24 | 0.34 |
| TP9 | -0.21 | 0.43 |
| TP10 | -0.08 | 0.78 |
| Delta Power |  |  |
| AF7 | 0.2 | 0.46 |
| AF8 | -0.32 | 0.22 |
| TP9 | -0.26 | 0.33 |
| TP10 | -0.11 | 0.68 |
| Theta Power |  |  |
| AF7 | -0.14 | 0.6 |
| AF8 | 0.01 | 0.97 |
| TP9 | -0.19 | 0.5 |
| TP10 | 0.44 | 0.11 |
| Alpha Power |  |  |
| AF7 | -0.51 | 0.07 |
| AF8 | 0.33 | 0.29 |
| TP9 | -0.52 | 0.09 |
| TP10 | -0.38 | 0.23 |
| Beta Power |  |  |
| AF7 | -0.43 | 0.16 |
| AF8 | -0.02 | 0.95 |
| TP9 | -0.49 | 0.11 |
| TP10 | -0.24 | 0.45 |
| Gamma Power |  |  |
| AF7 | -0.04 | 0.92 |
| AF8 | -0.28 | 0.44 |
| TP9 | -0.3 | 0.43 |
| TP10 | -0.52 | 0.19 |
| Full PLV  AF7, AF8 | 0.27 | 0.24 |
| AF7, TP9 | 0.35 | 0.13 |
| AF7, TP10 | 0.31 | 0.18 |
| AF8, TP9 | -0.14 | 0.57 |
| AF8, TP10 | -0.07 | 0.76 |
| TP9, TP10 | 0.05 | 0.83 |
| Delta  AF7, AF8 | **0.48** | **0.03** |
| AF7, TP9 | **0.55** | **0.01** |
| AF7, TP10 | 0.4 | 0.08 |
| AF8, TP9 | **0.56** | **0.01** |
| AF8, TP10 | 0.44 | 0.05 |
| TP9, TP10 | **0.49** | **0.03** |
| Theta  AF7, AF8 | 0.33 | 0.15 |
| AF7, TP9 | 0.34 | 0.14 |
| AF7, TP10 | 0.25 | 0.28 |
| AF8, TP9 | 0.35 | 0.13 |
| AF8, TP10 | 0.27 | 0.25 |
| TP9, TP10 | 0.42 | 0.07 |
| Alpha  AF7, AF8 | 0.33 | 0.16 |
| AF7, TP9 | 0.4 | 0.08 |
| AF7, TP10 | 0.24 | 0.31 |
| AF8, TP9 | 0.27 | 0.25 |
| AF8, TP10 | 0.28 | 0.24 |
| TP9, TP10 | 0.15 | 0.53 |
| Beta  AF7, AF8 | 0.32 | 0.17 |
| AF7, TP9 | 0.32 | 0.17 |
| AF7, TP10 | 0.16 | 0.5 |
| AF8, TP9 | 0.39 | 0.09 |
| AF8, TP10 | **0.45** | **0.04** |
| TP9, TP10 | 0.37 | 0.11 |
| Gamma  AF7, AF8 | 0.17 | 0.46 |
| AF7, TP9 | 0.24 | 0.32 |
| AF7, TP10 | 0.31 | 0.18 |
| AF8, TP9 | -0.25 | 0.29 |
| AF8, TP10 | -0.04 | 0.85 |
| TP9, TP10 | -0.28 | 0.24 |

*Note.* ^*^*p* < .05, ^**^*p* < .01, *p values are corrected for False Discovery Rate (FDR)*

**Supplementary Materials**

**Supplementary Table 6.**

Means, Standard Deviations, and main effect of Response Group (Remission/Non-Remission), main effect of time and interaction from Two-Way ANCOVA Analyses of Variance in Power and Phase Locking Value (PLV) and MADRS as covariate

| Variable | Main effect of Group | | | | | | Main effect of Time | | | | | | Interaction between group and time | |
| --- | --- | --- | --- | --- | --- | --- | --- | --- | --- | --- | --- | --- | --- | --- |
|  | Baseline | | Post-Treatment | | *F (*1,1) | *p* | Baseline | | Post-Treatment | | *F (*1,1) | *p* | *F (*1,1) | *p* |
|  | *M* | *SD* | *M* | *SD* |  |  | *M* | *SD* | *M* | *SD* |  |  |  |  |
| Full Power |  |  |  |  |  |  |  |  |  |  |  |  |  |  |
| AF7 | 979.59 | 1879.99 | 5896.89 | 13995.99 | 2.95 | .09 | 1744.76 | 4164.32 | 464.00 | 13039.17 | 1.03 | .32 | 1.92 | .17 |
| AF8 | 656.01 | 992.82 | 4211.90 | 1125.31 | 2.19 | .15 | 2088.16 | 6674.95 | 2424.17 | 8735.55 | .02 | .89 | 0.12 | .73 |
| TP9 | 4163.87 | 6199.73 | 3383.50 | 11134.19 | 0.08 | .78 | 1622.24 | 2371.90 | 6003.17 | 11747.06 | 2.52 | .12 | 0.12 | .73 |
| TP10 | 3111.23 | 6711.04 | 3225.81 | 10177.27 | 0.00 | .97 | 1082.47 | 1487.63 | 5243.11 | 11451.45 | 2.43 | .13 | 0.00 | .95 |
| Delta Power |  |  |  |  |  |  |  |  |  |  |  |  |  |  |
| AF7 | 225.74 | 324.61 | 2514.76 | 6732.03 | 2.87 | .10 | 318.43 | 581.20 | 2193.17 | 6419.40 | 1.94 | .17 | 2.08 | .16 |
| AF8 | 202.16 | 365.73 | 651.81 | 1916.94 | 1.28 | .27 | 232.72 | 417.55 | 576.29 | 1817.84 | .75 | .39 | 1.70 | .20 |
| TP9 | 606.63 | 1089.53 | 806.81 | 2901.12 | 0.09 | .76 | 427.39 | 1142.90 | 966.03 | 2721.13 | .68 | .42 | 1.55 | .22 |
| TP10 | 288.12 | 237.04 | 80.76 | 2763.88 | 0.79 | .38 | 184.15 | 144.84 | 853.47 | 2603.30 | 1.36 | .25 | 1.17 | .29 |
| Theta Power |  |  |  |  |  |  |  |  |  |  |  |  |  |  |
| AF7 | 61.65 | 69.33 | 533.04 | 1688.77 | 1.76 | .19 | 68.25 | 116.62 | 479.30 | 1603.21 | 1.36 | .25 | 1.48 | .23 |
| AF8 | 43.14 | 41.28 | 644.90 | 2526.67 | 1.32 | .26 | 47.64 | 54.44 | 58.21 | 2397.99 | 1.05 | .31 | 1.35 | .25 |
| TP9 | 193.32 | 263.71 | 891.43 | 3553.51 | 0.89 | .35 | 12.56 | 24.00 | 894.38 | 3355.04 | 1.11 | .30 | 1.31 | .26 |
| TP10 | 94.83 | 78.48 | 747.11 | 2917.47 | 1.16 | .29 | 67.52 | 53.38 | 709.20 | 2761.27 | 1.13 | .29 | 1.31 | .26 |
| Alpha Power |  |  |  |  |  |  |  |  |  |  |  |  |  |  |
| AF7 | 42.59 | 57.25 | 335.06 | 1093.17 | 1.61 | .21 | 52.05 | 97.42 | 296.35 | 1037.56 | 1.14 | .29 | 1.59 | .22 |
| AF8 | 28.40 | 25.59 | 336.40 | 1311.03 | 1.28 | .27 | 26.74 | 26.86 | 307.26 | 1243.34 | 1.07 | .31 | 1.38 | .25 |
| TP9 | 78.73 | 78.69 | 31.82 | 1148.13 | 0.94 | .34 | 56.48 | 7.39 | 309.87 | 1084.25 | 1.13 | .29 | 1.33 | .26 |
| TP10 | 51.78 | 33.78 | 535.70 | 2098.52 | 1.23 | .27 | 42.08 | 26.26 | 497.01 | 1988.44 | 1.10 | .30 | 1.31 | .26 |
| Beta Power |  |  |  |  |  |  |  |  |  |  |  |  |  |  |
| AF7 | 258.56 | 70.86 | 702.30 | 1623.54 | 1.51 | .23 | 431.80 | 1057.62 | 484.69 | 1372.55 | 0.02 | .88 | 1.29 | .26 |
| AF8 | 118.87 | 125.84 | 332.40 | 1025.01 | 0.97 | .33 | 123.50 | 158.09 | 306.42 | 968.87 | 0.72 | .40 | 1.35 | .25 |
| TP9 | 15.93 | 112.00 | 306.58 | 100.81 | 0.54 | .47 | 106.97 | 109.20 | 334.98 | 939.89 | 1.18 | .28 | 1.45 | .24 |
| TP10 | 123.19 | 101.42 | 288.41 | 923.51 | 0.72 | .40 | 95.97 | 92.99 | 299.11 | 87.81 | 1.10 | .30 | 1.54 | .22 |
| Gamma Power |  |  |  |  |  |  |  |  |  |  |  |  |  |  |
| AF7 | 401.30 | 1094.32 | 2191.19 | 6506.23 | 2.02 | .16 | 938.12 | 2613.45 | 1475.38 | 5821.17 | 0.18 | .67 | .85 | .36 |
| AF8 | 282.12 | 572.64 | 2335.89 | 7816.87 | 1.48 | .23 | 1891.79 | 7302.42 | 52.84 | 1707.94 | 0.67 | .42 | .55 | .46 |
| TP9 | 3531.54 | 6885.68 | 881.86 | 1771.02 | 2.62 | .11 | 986.15 | 1968.57 | 3692.22 | 7151.13 | 2.76 | .11 | 1.19 | .28 |
| TP10 | 2909.37 | 741.89 | 711.58 | 1714.81 | 1.56 | .22 | 765.94 | 1663.66 | 3074.78 | 7763.82 | 1.74 | .20 | 1.94 | .17 |
| Full PLV |  |  |  |  |  |  |  |  |  |  |  |  |  |  |
| AF7-AF8 | 0.29 | 0.16 | 0.19 | 0.19 | 3.42 | .07 | .28 | 0.23 | 0.20 | 0.11 | 2.17 | .15 | .18 | .68 |
| AF7-TP9 | 0.20 | 0.10 | 0.29 | 0.16 | **6.07** | **.02** | .21 | 0.12 | 0.27 | 0.15 | 2.62 | .11 | 1.91 | .18 |
| AF7-TP10 | 0.17 | 0.08 | 0.21 | 0.17 | 1.38 | .25 | .18 | 0.12 | 0.19 | 0.14 | 0.09 | .77 | 1.43 | .24 |
| AF8-TP9 | 0.17 | 0.14 | 0.18 | 0.14 | 0.02 | .88 | .14 | 0.09 | 0.20 | 0.17 | 2.24 | .14 | 0.00 | .99 |
| AF8-TP10 | 0.24 | 0.13 | 0.24 | 0.11 | 0.02 | .88 | .23 | 0.10 | 0.25 | 0.13 | 0.72 | .40 | 0.05 | .82 |
| TP9-TP10 | 0.60 | 0.25 | 0.52 | 0.22 | 1.14 | .29 | .53 | 0.24 | 0.59 | 0.24 | 0.83 | .37 | 0.15 | .70 |
| Delta PLV |  |  |  |  |  |  |  |  |  |  |  |  |  |  |
| AF7-AF8 | 0.38 | 0.15 | 0.26 | 0.09 | **8.65** | **.01** | .34 | 0.17 | 0.32 | 0.10 | 0.32 | .58 | 2.02 | .16 |
| AF7-TP9 | 0.35 | 0.13 | 0.31 | 0.04 | 1.99 | .17 | .34 | 0.13 | 0.33 | 0.08 | 0.14 | .71 | 1.75 | .19 |
| AF7-TP10 | 0.34 | 0.15 | 0.25 | 0.06 | **6.75** | **.01** | .32 | 0.15 | 0.28 | 0.10 | 0.74 | .40 | 0.71 | .40 |
| AF8-TP9 | 0.31 | 0.10 | 0.25 | 0.06 | 4.23 | .05 | .29 | 0.10 | 0.28 | 0.07 | 0.01 | .93 | 1.60 | .21 |
| AF8-TP10 | 0.34 | 0.11 | 0.32 | 0.07 | 0.71 | .41 | .34 | 0.11 | 0.33 | 0.08 | 0.14 | .71 | 0.50 | .48 |
| TP9-TP10 | 0.61 | 0.16 | 0.49 | 0.16 | **5.62** | **.02** | .57 | 0.18 | 0.54 | 0.15 | 0.35 | .56 | 1.48 | .23 |
| Theta PLV |  |  |  |  |  |  |  |  |  |  |  |  |  |  |
| AF7-AF8 | 0.32 | 0.16 | 0.18 | 0.06 | **11.59** | **.00** | .28 | 0.17 | 0.25 | 0.11 | 0.59 | .45 | 0.72 | .40 |
| AF7-TP9 | 0.34 | 0.14 | 0.29 | 0.07 | 1.77 | .19 | .32 | 0.13 | 0.32 | 0.11 | 0.00 | .97 | 0.59 | .45 |
| AF7-TP10 | 0.26 | 0.14 | 0.15 | 0.03 | **8.80** | **.01** | .22 | 0.14 | 0.21 | 0.09 | 0.07 | .79 | 0.51 | .48 |
| AF8-TP9 | 0.23 | 0.10 | 0.19 | 0.07 | 2.27 | .14 | .21 | 0.09 | 0.22 | 0.09 | 0.11 | .75 | 0.84 | .37 |
| AF8-TP10 | 0.32 | 0.12 | 0.32 | 0.07 | 0.01 | .94 | .32 | 0.11 | 0.32 | 0.09 | 0.00 | .95 | 0.00 | .99 |
| TP9-TP10 | 0.55 | 0.14 | 0.42 | 0.15 | **7.92** | **.01** | .51 | 0.17 | 0.48 | 0.14 | 0.33 | .57 | 1.55 | .22 |
| Alpha PLV |  |  |  |  |  |  |  |  |  |  |  |  |  |  |
| AF7-AF8 | 0.31 | 0.17 | 0.18 | 0.07 | **9.57** | **.00** | .26 | 0.16 | 0.24 | 0.14 | 0.34 | .56 | 0.71 | .40 |
| AF7-TP9 | 0.39 | 0.13 | 0.31 | 0.08 | **5.69** | **.02** | .34 | 0.14 | 0.36 | 0.08 | 0.35 | .56 | 0.47 | .50 |
| AF7-TP10 | 0.25 | 0.12 | 0.16 | 0.04 | **8.33** | **.01** | .21 | 0.12 | 0.21 | 0.08 | 0.00 | .98 | 0.48 | .49 |
| AF8-TP9 | 0.25 | 0.11 | 0.19 | 0.06 | 3.55 | .07 | .22 | 0.10 | 0.22 | 0.09 | 0.03 | .86 | 0.32 | .58 |
| AF8-TP10 | 0.38 | 0.11 | 0.34 | 0.09 | 1.59 | .22 | .37 | 0.10 | 0.35 | 0.10 | 0.41 | .53 | 0.02 | .90 |
| TP9-TP10 | 0.53 | 0.13 | 0.42 | 0.09 | **7.87** | **.01** | .49 | 0.14 | 0.47 | 0.11 | 0.33 | .57 | 0.05 | .82 |
| Beta PLV |  |  |  |  |  |  |  |  |  |  |  |  |  |  |
| AF7-AF8 | 0.29 | 0.15 | 0.21 | 0.17 | 2.75 | .11 | .28 | 0.19 | 0.23 | 0.13 | 0.75 | .39 | 0.33 | .57 |
| AF7-TP9 | 0.34 | 0.11 | 0.24 | 0.12 | **9.49** | **.00** | .30 | 0.14 | 0.29 | 0.11 | 0.01 | .94 | 0.92 | .34 |
| AF7-TP10 | 0.25 | 0.10 | 0.17 | 0.11 | **7.03** | **.01** | .23 | 0.12 | 0.20 | 0.10 | 0.53 | .47 | 1.00 | .32 |
| AF8-TP9 | 0.22 | 0.11 | 0.19 | 0.10 | .72 | .40 | .21 | 0.11 | 0.21 | 0.10 | 0.03 | .86 | 2.28 | .14 |
| AF8-TP10 | 0.32 | 0.09 | 0.28 | 0.10 | 1.64 | .21 | .31 | 0.08 | 0.29 | 0.11 | 0.26 | .61 | 2.39 | .13 |
| TP9-TP10 | 0.38 | 0.10 | 0.35 | 0.10 | .97 | .33 | .36 | 0.10 | 0.38 | 0.10 | 0.79 | .38 | **4.37** | **.04** |
| Gamma PLV |  |  |  |  |  |  |  |  |  |  |  |  |  |  |
| AF7-AF8 | 0.34 | 0.17 | 0.26 | 0.21 | 1.42 | .24 | .32 | 0.23 | 0.29 | 0.14 | 0.34 | .56 | 0.03 | .87 |
| AF7-TP9 | 0.27 | 0.16 | 0.39 | 0.29 | 3.53 | .07 | .28 | 0.20 | 0.37 | 0.25 | 2.08 | .16 | 0.10 | .76 |
| AF7-TP10 | 0.24 | 0.17 | 0.36 | 0.27 | 3.60 | .07 | .28 | 0.19 | 0.32 | 0.26 | 0.48 | .49 | 0.33 | .57 |
| AF8-TP9 | 0.29 | 0.22 | 0.28 | 0.21 | 0.08 | .78 | .21 | 0.16 | 0.37 | 0.23 | **6.65** | **.01** | 0.28 | .60 |
| AF8-TP10 | 0.33 | 0.23 | 0.29 | 0.16 | 0.44 | .51 | .27 | 0.17 | 0.35 | 0.23 | 1.69 | .20 | 0.03 | .86 |
| TP9-TP10 | 0.63 | 0.35 | 0.60 | 0.27 | 0.08 | .77 | .51 | 0.32 | 0.72 | 0.27 | **5.68** | **.02** | 0.52 | .47 |

*Note. ^*^p < .05, ^**^p < .01, p values are corrected for False Discovery Rate (FDR)*
